# Understanding Clinical Reasoning Variability in Medical Large Language Models: A Mechanistic Interpretability Study

**DOI:** 10.64898/2026.01.26.26344845

**Authors:** Mirage Modi, Jordan E. Krull, Donte Johnson, Xiaoying Wang, Timothy D. Gauntner, Mingjia Li, Hao Cheng, Anjun Ma, Ping Zhang, Daniel G. Stover, Zihai Li, Qin Ma

## Abstract

Medical large language models (LLMs) achieving high benchmark accuracy exhibit unexplained variability in clinical tasks, producing errors that clinicians cannot safeguard against. We evaluated clinical reasoning stability in GPT-5, MedGemma-27B-Text-IT, and OpenBioLLM-Llama3-70B using 355 systematic perturbations of physician-validated oncology cases and trained sparse autoencoders on 1 billion tokens from 50,000 MIMIC-IV clinical notes to decompose their internal representation. We find models exhibit dramatic reasoning instability, shifting staging accuracy by over 50% based solely on prompt format, or generating definitive staging in clinically insufficient scenarios. Sparse autoencoder analysis revealed hierarchical encoding in MedGemma, where high-magnitude features encode lexical identity and low-magnitude features encode contextual meaning. OpenBioLLM distributes information uniformly. We demonstrate these internal encoding structures differentially affect retrieval interventions, suggesting interventions effective for one architecture may harm another. We recommend healthcare institutions implement architecture-specific safety validation, as benchmark equivalence does not imply functional equivalence, with implications for AI safety beyond healthcare.

---

Large language models (LLMs) are rapidly entering clinical practice. Medical question-answering systems now report over eight million queries monthly, and domain-specific models achieve near-expert performance on standardized medical examinations.^1–3^ Med-PaLM 2 attains 86.5% accuracy on United States Medical Licensing Examination–style questions, with physician evaluators preferring its responses over those written by clinicians on most quality dimensions.^2^ More recent open-source medical models, including MedGemma and OpenBioLLM, demonstrate comparable benchmark performance while offering greater accessibility for research and deployment.^3,4^ These advances have accelerated enthusiasm for integrating LLMs, as clinical decision support tools, into clinical documentation, diagnostic support, and treatment planning.

However, benchmark performance provides an incomplete and potentially misleading picture of clinical readiness. Recent evaluations reveal that models achieving superhuman scores on multiple-choice examinations exhibit striking deficits in clinical reasoning under realistic conditions. Beyond accuracy metrics, LLMs demonstrate fragile clinical reasoning, showing systematic overconfidence on diagnostic belief revision tasks, sensitivity to minor prompt wording changes (up to 10% accuracy shifts), and susceptibility to propagating planted false information in up to 83% of adversarial cases.^5–7^

Current evaluation paradigms rely on accuracy metrics from curated benchmarks, which fail to distinguish whether models encode clinical knowledge through robust conceptual representations or through superficial statistical regularities. This distinction matters because models achieving similar benchmark performance may differ fundamentally in their capacity for reliable generalization. Yet, current accuracy and safety interventions do not reliably alter model behavior.^8,9^ We hypothesize that mechanistic interpretability methods, specifically sparse autoencoders applied to model internals, can reveal whether medical LLMs develop stable, clinically meaningful representations, or whether their apparent competence masks fragile reasoning that falters under perturbation.

Rather than treating models as black boxes evaluated solely by their predictions, mechanistic methods aim to identify the computational features and circuits through which models process information.^10^ Sparse autoencoders (SAEs) have emerged as an effective technique for decomposing neural network activations into interpretable components.^11^ They learn dictionaries of features, corresponding to human-interpretable concepts. When applied to general-domain LLMs, SAEs have revealed features encoding concepts ranging from protein sequences and legal language to abstract notions of deception and sycophancy.^12–14^

Open resources, including Gemma Scope, provide pretrained autoencoders for the Gemma model family, encompassing over 400 autoencoders with more than 30 million learned features.^15^ Yet despite this methodological maturity and the clear relevance to clinical artificial intelligence governance, no published work has systematically applied SAEs for the mechanistic interpretability of medical LLMs. This gap is consequential: healthcare applications face stringent requirements for explainability, with regulatory guidance indicating that clinical decision support lacking interpretable mechanisms may require device-level oversight.

In this study, we sought to address this gap by (i) characterizing the medical-LLM critical reasoning variability when evaluating complex cases, (ii) elucidating mechanisms of poor reasoning structure in realistic scenarios, and (iii) intervening when models exhibit behavioral instability. Through prompt reasoning tests on realistic oncology cases, we identify weak points in prompt structure and trait attribution for accurate assessment. We then evaluate context and reasoning structures within two open-source, SOTA medical LLMs (MedGemma-27B-Text-IT and OpenBioLLM-Llama3-70B)^3,4,16^ by training and systematically analyzing large-scale SAEs on model activations of clinical notes sampled from Medical Information Mart for Intensive Care (MIMIC-IV-Note), a corpus of over 330,000 deidentified discharge summaries representing diverse critical care encounters.^17,18^ With these methods, we find that both LLMs use diverse features to encode medical knowledge, that context-encoding features are causally necessary for clinical prediction, and that clinical safety interventions require architecture-awareness to improve clinical understanding.

## Results

### Benchmark performance does not predict clinical reasoning stability

Benchmarking tasks in LLMs have largely involved question-and-answer on relatively routine medical situations, so we evaluated whether medical AI systems maintain consistent clinical reasoning under realistic conditions by presenting three synthetic oncology cases to leading medical LLMs: GPT-5 Thinking, MedGemma-27B-Text-IT, and OpenBioLLM-Llama3-70B. Each case was constructed in a standard SOAP format by two board-certified medical oncology physicians, to mimic a real scenario seen in an outpatient oncology clinic visit. We systematically removed individual medical terms (see methods) from each case, yielding 355 total perturbation trials per model (**Figure 1a**). We prompted several models to produce an A&P from the S&O, which displayed marked variability compared to physician-produced A&P, when no medical terms were perturbed (**Figure 1b**). Using two open source medical LLMs and GPT-5 Thinking, each model was prompted iteratively for each perturbed version of the notes to produce an A&P, under two prompt formats (produce a full A&P or direct prompting for clinical stage and treatment), which displayed marked variability between models in tumor staging, depending on prompt format (**Figure 1c**). Staging accuracy varied substantially with prompt structure in model- and case-specific manners. For UC, OpenBioLLM accuracy shifted from 45.9% to 99.1% based solely on whether staging was embedded in a full A&P or queried directly; MedGemma showed similar instability (74.8% to 98.2%). GPT-5 Thinking was largely incorrect on NSCLC-IT (93-100% incorrect). Most concerning, on NSCLC-FU, where clinical information was intentionally insufficient, MedGemma provided definitive staging 100% of the time when directly prompted, whereas GPT-5 appropriately indicated uncertainty. This inconsistent reliability across models and cases undermines clinical trust regardless of any single high-accuracy result.

**Figure 1.**
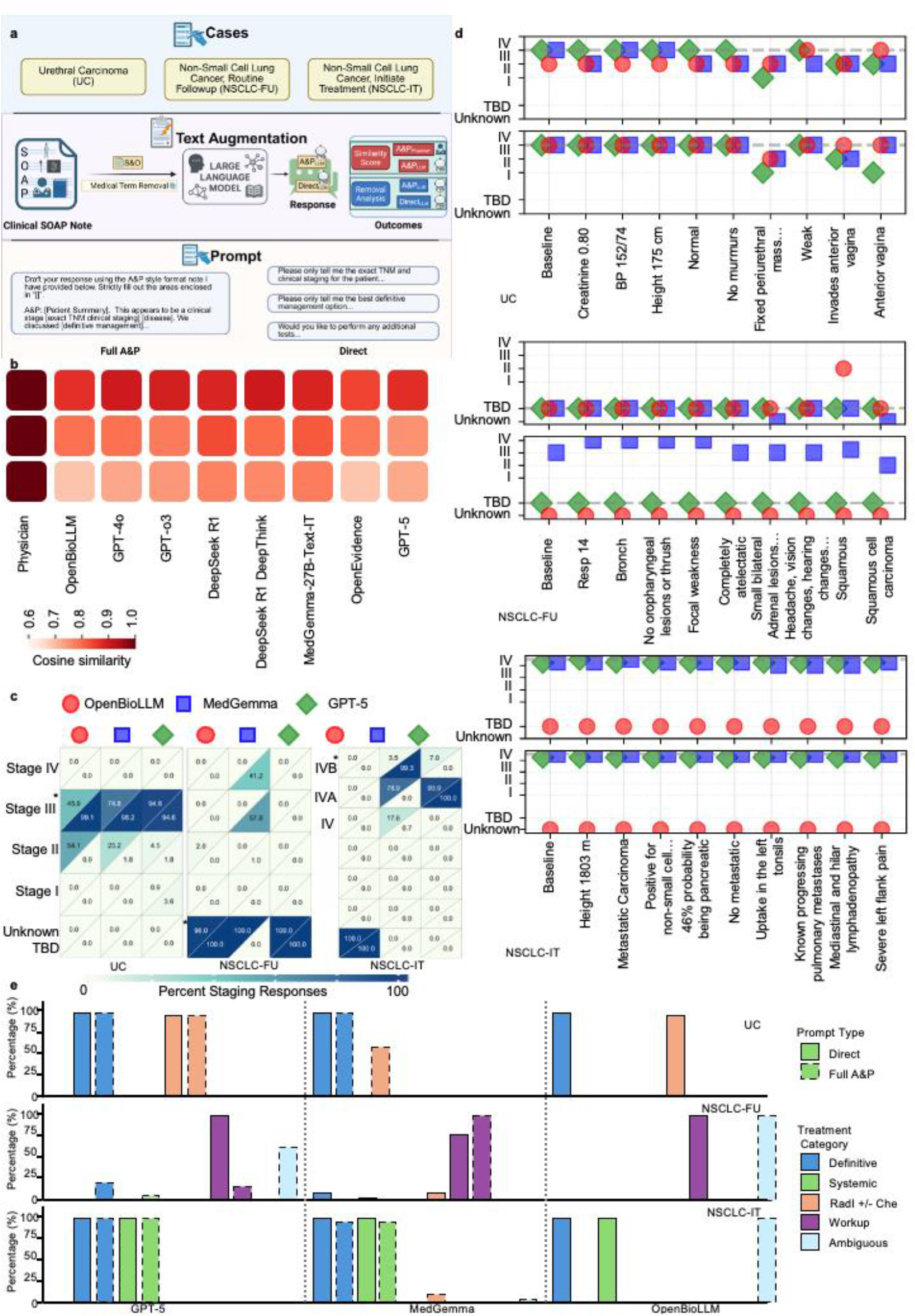
Text augmentation leads to unstable LLM clinical reasoning and decision-making. (a) Three realistic oncology cases—Urethral Carcinoma (UC), a clinical follow-up for NSCLC (NSCLC-FU), and an initial treatment visit for Non-Small Cell Lung Cancer (NSCLC-IT)—were tested as SOAP notes, with Subjective & Objective (S&O) serving as input, with medical terms being removed and different prompt strategies applied. (b) Cosine similarity scores comparing text embeddings from LLM Assessment & Plan (A&P) outputs compared to the physician A&P embedding when no terms are removed, with a higher score indicating greater similarity to the physician note. BioLORD-2023 is used to produce the embeddings. (c) LLM output tumor staging separated by model (columns: GPT-5 Thinking, OpenBioLLM, and MedGemma) and full-A&P vs. Direct Prompt (upper left vs. lower right), the heatmap represents the percentage of input terms that yield that stage when deleted from the prompt, with the * indicating the correct tumor staging. (d) Selected terms from the input prompt and the model’s clinical stage output when the term or phrase is removed from the prompt, separated by full A&P prompt or direct prompt strategies. Green diamond – GPT5 Thinking, blue square – MedGemma, orange circle – OpenBioLLM, dotted line – ground truth. (e) Percentage of output predictions of treatment category (definitive management, systemic therapy, radiation +/-chemotherapy, workup, and ambiguous treatment options) for all terms deleted from the prompt, split by model and full A&P vs. direct prompt.

MedGemma and OpenBioLLM exhibited sensitivity to clinically irrelevant information (**Figure 1d**). Removing “creatinine 0.80” from the UC prompt, a term with no relevance to tumor staging, resulted in MedGemma changing its staging from correct (Stage III) to incorrect Stage II. Conversely, when clinically relevant terms such as “invades the anterior vagina” were removed, OpenBioLLM maintained its original staging, with similar staging variability for MedGemma direct prompting when staging is unknown (NSCLC-FU). GPT-5 demonstrates resistance to irrelevant text perturbations and correctly changes its answers in a clinically aligned manner for relevant terms (“anterior vagina”, UC) but also remains consistent even when incorrect (NSCLC-IT).

Treatment recommendations showed comparable variability (**Figure 1e**). For NSCLC-IT, OpenBioLLM recommended definitive treatment options in 99.3% of full A&P responses but provided an ambiguous treatment recommendation in 100% of direct queries. When staging is unknown, we find the models tend to provide a spread of treatment recommendations (NSCLC-FU). These findings indicate that benchmark performance, where all tested models achieve state-of-the-art accuracy on standardized medical examinations, fails to predict reasoning stability in clinical contexts.

### Chain-of-thought prompting does not resolve reasoning deficits

Because the models provided more treatment recommendations when staging was unknown, we next tested whether explicitly prompting models to reason stepwise through TNM staging before providing treatment recommendations would improve consistency (**Figure 2a**). Under chain-of-thought (CoT) prompting, models first extracted TNM components, then derived clinical stage, then recommended treatment, mimicking clinical reasoning workflows. CoT prompting did not reliably improve staging accuracy. When models’ TNM-derived clinical stages were compared against AJCC 9^th^ edition staging guidelines (or 8^th^ edition for UC) across all 355 perturbations, systematic errors persisted. GPT-5 achieved 97.5% concordance, while MedGemma achieved only 26.8% and OpenBioLLM achieved 31.0% (**Figure 2b**). For example, MedGemma reported “T4N3M1b (Stage IVB)”, which should be Stage IVA according to the AJCC 9^th^ edition. This pattern suggests memorized associations rather than principled applications of staging algorithms.

**Figure 2.**
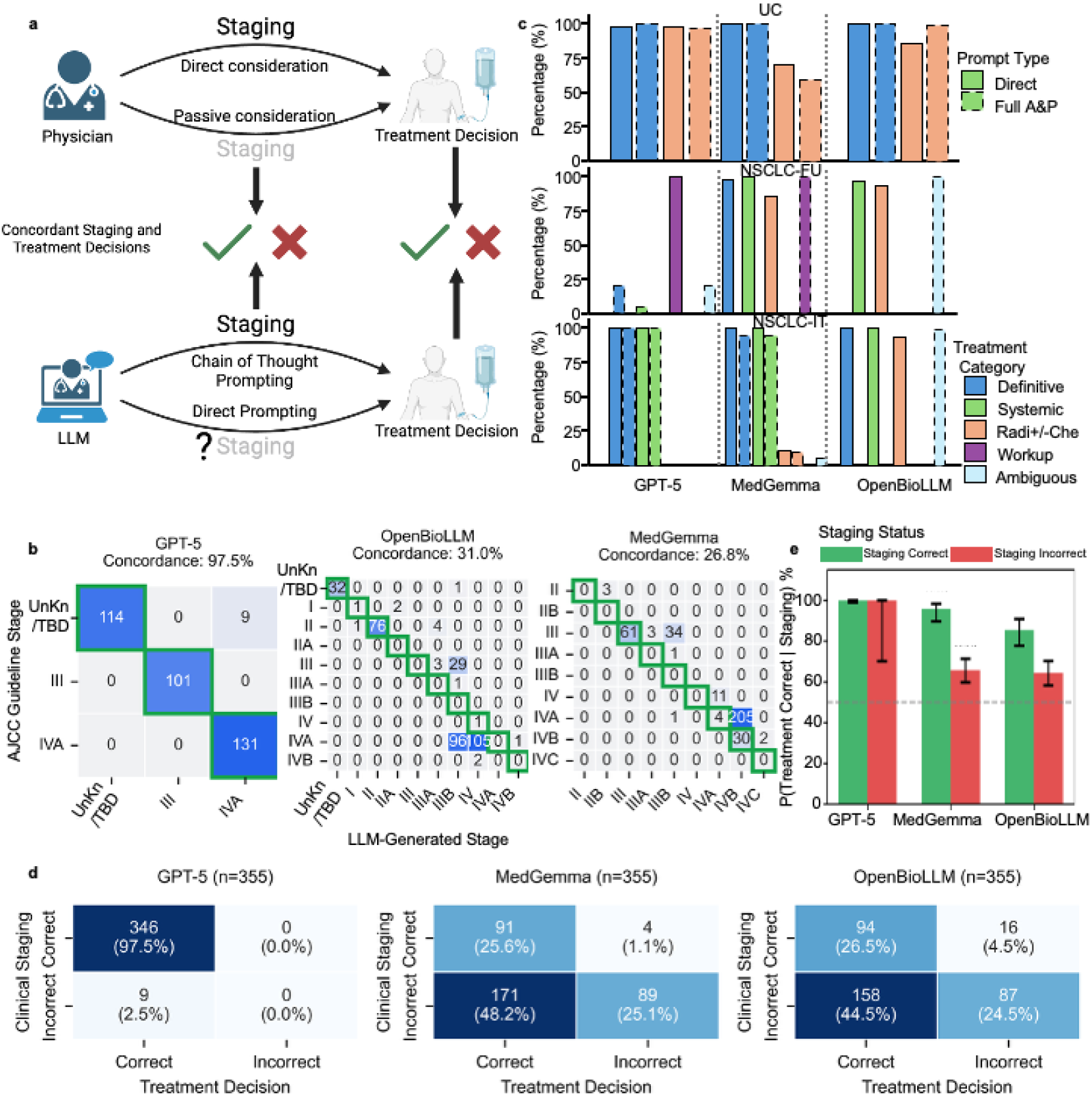
Chain-of-thought prompting reveals LLM memory mapping using tumor staging. (a) Schematic of the chain-of-though (CoT) evaluation: models first extract TNM staging, then derive clinical stage, then recommend treatment, compared against direct treatment queries to measure concordance to a physician’s clinical thinking. (b) Confusion matrices comparing LLM-derived clinical staging (columns) against AJCC 8/9^th^ edition staging guidelines (rows) for each model and case. Overall AJCC staging concordance was 97.5% for GPT-5, 26.8% for MedGemma, and 31.0% for OpenBioLLM. (c) Treatment category proportions by model and prompting strategy (CoT versus Direct) across three oncology cases (NSCLC-FU, NSCLC-IT, UC). Treatment categories include definitive management, systemic therapy, radiation +/-chemotherapy, and ambiguous treatment options, defined by NCCN treatment guidelines. Notable findings include OpenBioLLM shifting from 99.3% definitive to 100% ambiguous for NSCLC-IT between CoT and Direct prompting, and MedGemma recommending definitive treatment in 98.0% of NSCLC-FU CoT responses despite staging uncertainty. (d) Confusion matrix of LLM clinical staging versus LLM treatment decision. MedGemma correctly staged 97.5% of responses with correct downstream treatment. MedGemma and OpenBioLLM correctly staged only 25.6% and 26.5% of responses, respectively, with correct downstreatm treatment. (e) Correct treatment dependency on correct stage. GPT-5 gave correct treatments 100% of the time, making it independent of correct staging (p=1.0, Fisher’s exact test). MedGemma and OpenBioLLM are dependent on correct staging (p<0.001, Fisher’s exact test).

Treatment concordance between CoT and direct prompting strategies varied substantially by case complexity (**Figure 2c**). For the UC case, all models recommended appropriate definitive management regardless of prompting strategy. However, for NSCLC-FU (staging to be determined), MedGemma recommended definitive management in 98% of CoT responses, while GPT-5 appropriately recommended workup in 100% of responses. OpenBioLLM recommended radiation or systemic therapy in over 90% of responses without addressing the staging ambiguity. GPT-5 provided correct treatment independent of correct staging (**Figure 2d-e**), while MedGemma and OpenBioLLM both display a dependence on staging to arrive at the correct treatment (p<0.001, Fisher’s exact test).

### Sparse autoencoder analysis reveals interpretable clinical concept organization

To understand why models fail at clinical reasoning despite benchmarking success, we trained top-k sparse autoencoders (SAE) to decompose model activations into interpretable components (**Figure 3a**). Dimensionality reduction on MedGemma’s SAE revealed that learned features cluster into coherent clinical concept groups (**Figure 3b**). HDBSCAN clustering identified five distinct groupings corresponding to medication lists for cardiopulmonary patients, complex medicine inpatient cases, neurovascular procedures, alcohol-related adverse events, and general medical documentation. Local lexical coherence analysis demonstrated that tokens with high feature overlap cluster spatially (**Figure 3c**), indicating non-random organization of clinical concepts in feature space. The highest effect sizes were seen among the clusters associated with complex medicine inpatient cases (fever in immunocompromised, respiratory decompensation), suggesting an association between clinically complex cases and reliance on token frequency.

**Figure 3.**
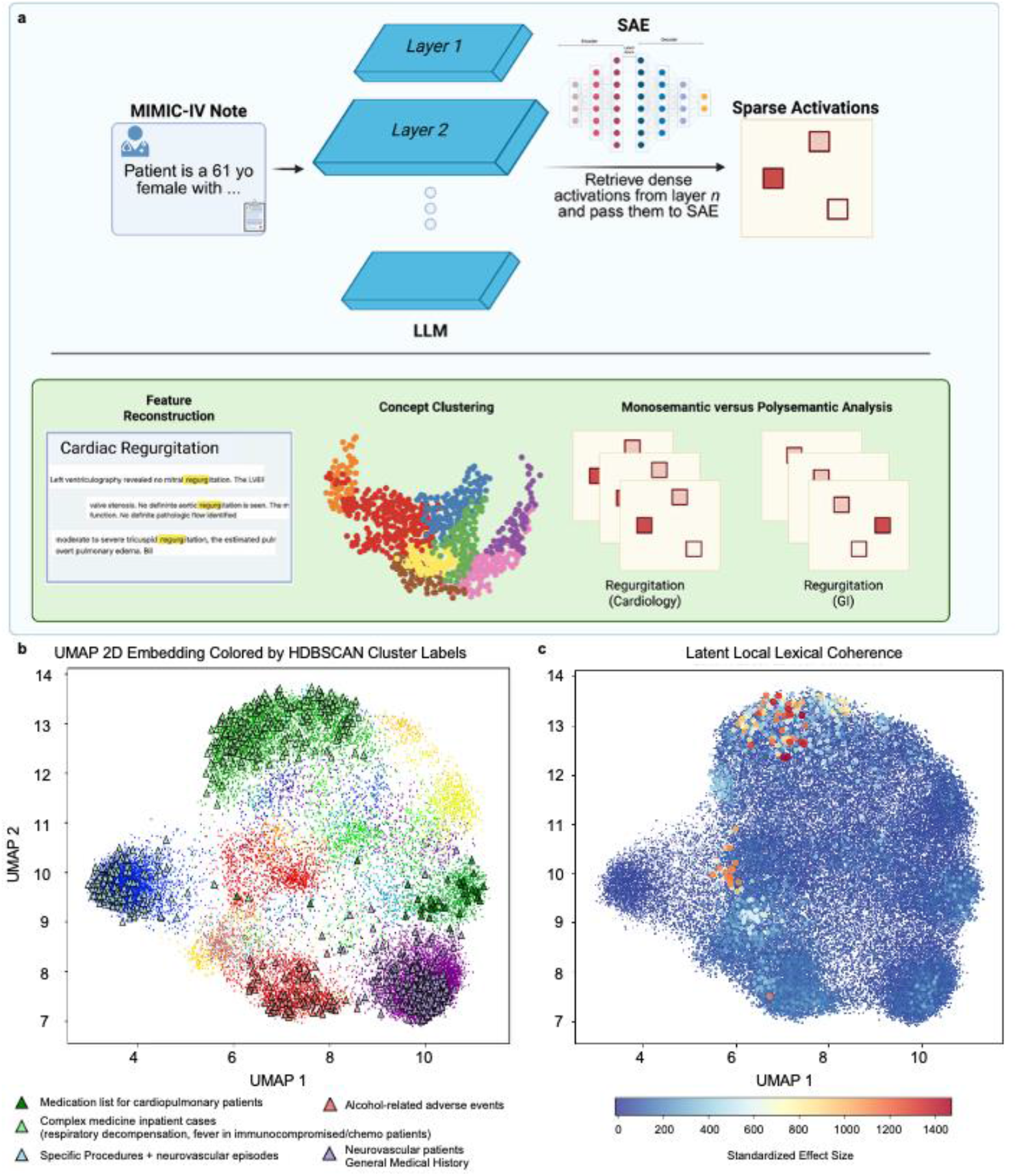
Sparse autoencoder reveals medical concepts in MedGemma-27B-Text-IT. (a) Graphical representation of the sparse autoencoder training. MIMIC-IV notes are sent to the medical LLM, retrieving dense activations from an internal layer. The sparse autoencoder (SAE) learns a sparse representation of the activations, from which we then perform downstream analyses. (b) A UMAP embedding of SAE latents clustered on their dictionary values using HDBScan, with the cluster color based on HDBScan cluster labels. Diamond-shaped points are randomly selected latents from 5 of the largest latent clusters, which share conceptual value. Each point is represented by a concept listed below in the legend, which summarizes the top terms for all latents in a cluster. (c) A UMAP embedding of SAE latents, with scaled values for the standardized effect size of local lexical coherence among top terms of each latent, represented by both point color and size. Large values represent more local overlap of top latent terms, above expected word frequency, and small values represent less overlap among top latent terms within each latent’s local neighborhood, than the expected term frequency overlap. Aggregation of higher value points shows local concept dependence on the presence of specific terms above the expected random occurrence frequency.

Qualitative examination confirmed clinical interpretability (**Table 1**). Feature 418 was activated maximally for dental terminology. Feature 56 bundled echocardiogram terminology (“size”, “thickness”, “motion”, “diameter”), co-activated due to their textual proximity in the clinical notes, illustrating how next-token prediction objectives shape feature organization. Feature 52, associated with lexical meanings of household furniture (“pilar”, “chairs”, “mattress”), instead activated for gastrointestinal content (“stool softener”, “stool extended constipation”), demonstrating unexpected clinical semantics.

**Table 1.** Representative sparse autoencoder latents demonstrate interpretable clinical concept encoding. Selected latents from MedGemma illustrating the relationship between lexical neighborhoods (top tokens by activation magnitude) and clinical semantics (top-activating text examples). Latent 418 encodes dentistry-specific terminology. Latent 538 captures neurological gait descriptors used in physical examination documentation. Latent 56 bundles echocardiogram terminology containing semantically distinct terms (“size”, “motion”) co-activated due to textual proximity in clinical notes, illustrating how MedGemma’s next-token prediction objectives shape feature organization. Latent 53 demonstrates unexpected lexical-semantic mapping, in which tokens associated with furniture (“pillar”, “chairs”, “mattress”) instead activate for gastrointestinal clinical content (“stool softener”, “stool extended constipation”).

| Latent | Top Tokens | Top Examples | Interpretation |
| --- | --- | --- | --- |
| 418 | periodontal;<br>tooth; dental;<br>molar; fitting;<br>denture;<br>amalgam;<br>wisdom; caries | "Pt presents with heavily restored dentition with multiple missing teeth. The upper molar as noted on the image as left side shows evidence of periapical pathology"<br>"DENTITION: Multiple maxillary teeth are absent. There is a periapical lucency in the location tooth #6 and #7 (3: 84)"<br>"Chronic Fatigue Syndrome Endocarditis, 1980s following dental procedure" | Dentistry-specific findings with lexical meaning related to dental materials and pathology |
| 538 | Steady; steadily;<br>speedy; slow;<br>deliberate;<br>leisurely;<br>stubbornly | "cerebellar exam intact, steady gait"<br>"Exam was notable for a steady gait"<br>"sensation grossly intact throughout, steady gait" | Neurological exam findings on gait with lexical meanings related to gait descriptors |
| 56 | Size; thickness;<br>motion; height;<br>circumference;<br>diameter | "the left ventricular cavity size is normal. Regional left ventricular wall motion is normal."<br>"The left ventricular cavity size is normal. There is moderate global left ventricular hypokinesis"<br>"Right ventricular chamber size and free wall motion are normal." | Echocardiogram descriptions of heart chambers with lexical meaning of size/dimension except motion is a distinct lexical meaning; these concepts are bundled on textual proximity since "size" and "motion" are collocated neighbors. |
| 53 | Pillar; chairs;<br>mattress | "stool softener, NOT a laxative..."<br>"stool extended constipation..."<br>"stool softeners, and should eat foods..." | GI disruption examples with lexical meaning of household furniture items |

### Architecturally distinct models encode medical polysemy differently

Medical terminology frequently carries multiple meanings depending on context. We examined 12 polysemous terms and 4 monosemantic terms (**Tables S2-3**) to determine whether models reliably distinguish contextual meanings. MedGemma exhibited a distinctive two-tier organization (**Figure 4a**). The top 10 highest-magnitude features showed 77.8% ± 8.7% overlap for polysemous terms (n=182 samples) and 78.8% ± 4.1% for monosemantic terms (n=2,392 samples). However, when examining all 128 active features, overlap dropped to 16.3% ± 0.3% for polysemous terms (n=183 samples) and 17.0% ± 4.9% for monosemantic terms (n=20,000 comparisons). This 4.6-fold reduction indicates that low-magnitude features encode contextual meaning rather than word identity. OpenBioLLM showed no such separation. Top-10 polysemantic feature overlap was 13.6% ± 3.1% (n=320 samples) and 17.9% ± 2.1% for monosemantic terms (n=2,661 samples), similar to the full 128-latent overlap of 10.3% ± 0.7% (polysemous, n=317 samples) and 11.6% ± 6.2% (monosemantic, n=20,000 samples). This finding suggests information is distributed uniformly across all features regardless of magnitude. The difference between architectures in top-10 polysemous overlap was significant (p<0.001, two-sample Z-test). Notably, OpenBioLLM showed significantly lower k=10 overlap for polysemous versus monosemantic terms (p<0.001, empirical distribution test), while MedGemma showed no such difference (p=0.75).

**Figure 4.**
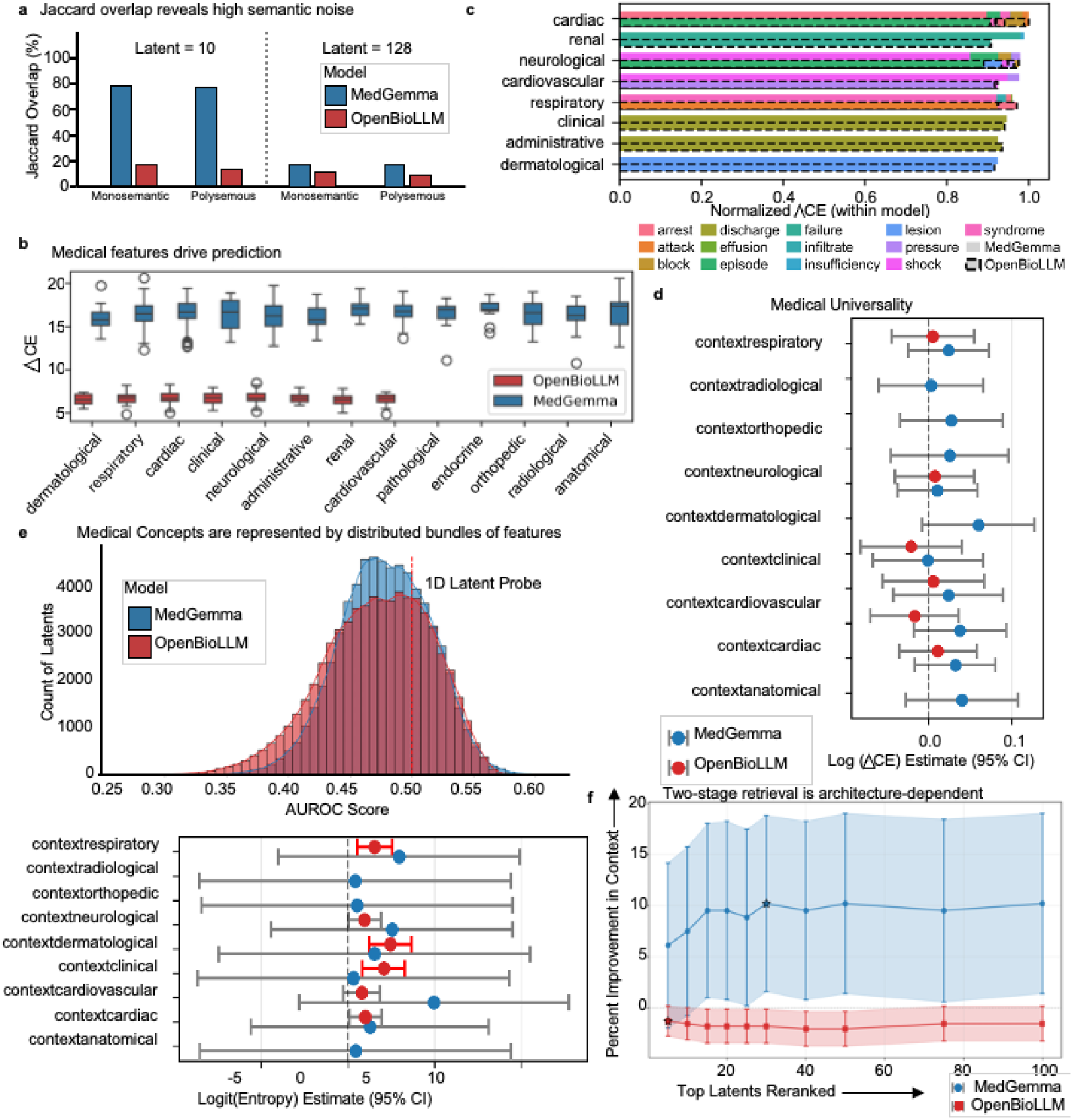
Sparse autoencoder analysis reveals architecture-specific encoding of medical polysemy. (A) Jaccard overlap between feature sets activated for polysemous versus monosemantic medical terms at k=10 (top features only) and k=128 (all active features). MedGemma exhibits 77.8% overlap at k=10 regardless of word sense, dropping to 16.9% at k=128, indicating hierarchical separation of lexical identity (high-magnitude features) from medical contextual meaning (low-magnitude features). OpenBioLLM shows uniformly low overlap (10-18%) at both k values, indicating distributed clinical representations. The between-architecture difference at k=10 (64.2%) was significant (p < 0.001). (B) Change in cross-entropy loss (Δ*CE*) when ablating context-encoding features, grouped by clinical sense. MedGemma ablations produced mean Δ*CE* of 16.43 ± 1.59 nats (n=445 ablations) versus 6.69 ± 0.66 nats for OpenBioLLM (n=400 ablations). (C) The lemmas with highest ablation impact by sense. (D) Mixed-effects model coefficients for log(Δ*CE*) by clinical context. Neither model showed significant context-dependent ablation effects (p > 0.05, gamma mixed effects), indicating domain-general mechanisms. (E) Distribution of single-latent probe AUROC scores for distinguishing clinical entities from background text and mixed-effects model coefficients for normalized feature entropy by clinical context.. Mean AUROC was 0.479 for MedGemma and 0.472 for OpenBioLLM, both indistinguishable from chance, with no probe exceeding 0.70. MedGemma showed no significant context effects (all p > 0.05, beta mixed effects); OpenBioLLM showed elevated entropy in dermatological (p = 0.002, beta mixed effects), clinical (p = 0.007, beta mixed effects), and respiratory (p = 0.011, beta mixed effects) contexts. (F) Two-stage retrieval schematic and results. Standard retrieval ranks by full feature similarity; two-stage retrieval first retrieves candidates, then re-ranks using only semantic features (positions 11–128). MedGemma showed gains of 10.2% in precision at rank 1 (p = 0.022, n=147 queries, paired t-test); OpenBioLLM showed loss of 2.0% (p = 0.021, n=395 queries, paired t-test). Stars indicate optimal k values.

### Context-encoding features are causally necessary for clinical prediction and are represented by distributed bundles of features

We tested whether context-specific features are functionally necessary for the LLM through targeted ablation experiments (**Figure 4b**). For each of the 12 polysemous terms, we identified features associated with specific clinical meanings via co-occurrence statistics and “disabled” them, measuring the change in cross-entropy loss (Δ*CE*; **Equation 1**) for next-token prediction. Disabling context-encoding features in MedGemma produced severe prediction degradation (mean Δ*CE* =16.43 ± 1.59 nats, n=445 ablations), representing near-complete loss of predictive capability. Effects were consistent across clinical domains: cardiovascular (16.64±1.25 nats, n=31 ablations), renal (17.02±1.13 nats, n=29 ablations), and administrative (16.02±1.47 nats, n=19 ablations). OpenBioLLM showed qualitatively similar but smaller effects (mean Δ*CE* =6.69

± 0.66 nats, n=400 ablations). The top senses show the specific lemma and its contribution to the Δ*CE* **(Figure 4c)**. Mixed effects regression with random intercepts for clinical notes confirmed these effects were domain-general (**Figure 4d**). For MedGemma and OpenBioLLM, no clinical context showed significant deviation from the overall mean (p>0.05 for all domains, gamma mixed effects), thus disabling context-specific features produced statistically equivalent prediction degradation within each model.

We then tested whether individual features encode broad medical significance by training univariate logistic regression probes per latent to distinguish clinical entities from background text (65,536 probes for MedGemma; 98,304 probes for OpenBioLLM; **Figure 4e**). Mean AUROC was 0.479±0.037 for MedGemma and 0.472±0.044 for OpenBioLLM, both indistinguishable from chance (0.50), with no probe achieving an AUROC above 0.70. No single feature reliably identifies medical content, meaning medical knowledge is distributed across thousands of specialized features. Mixed-effects analysis of normalized feature entropy revealed divergent architecture-specific patterns. MedGemma showed uniformly high entropy across all clinical contexts with no significant context effects (p>0.05, beta mixed effects). In contrast, OpenBioLLM showed significant context-dependent variation. Dermatological contexts exhibited elevated entropy relative to baseline (coefficient 2.46, 95% CI 1.23-3.68, p =0.002, beta mixed effects), followed by clinical contexts (coefficient 2.06, 95% CI 0.83-3.28, p = 0.007) and respiratory contexts (coefficient 1.54, 95% CI 0.54-2.55, p=0.011). This suggests that while MedGemma maintains consistent representational complexity across specialties, OpenBioLLM develops domain-specific representational patterns with greater feature dispersion in certain clinical contexts.

### Architecture-aware retrieval improves disambiguation for MedGemma but harms for OpenBioLLM

The architectural differences we identified suggest a practical intervention. If MedGemma concentrates word identity in high-magnitude features and contextual meaning in low-magnitude features, retrieval systems could improve disambiguation by filtering identity-encoding features when re-ranking. We tested this hypothesis using a same-word sense retrieval task with terms meeting minimum thresholds of 30 total instances and at least 2 senses with 5 or more instances each (**Figure 4f**). We employ a two-stage retrieval where we retrieve top-k candidates using all 128 features, then re-rank using only features 11-128. For MedGemma (n=147 queries across 8 polysemous terms), two-stage retrieval produced a significant improvement. At optimal k=30, precision at rank one improved by 10.2% (95% CI: 1.6-18.8; p=0.022, paired t-test). This same intervention harmed OpenBioLLM (n=396 queries across 10 polysemous terms). At k=40, precision decreased by 2.0% (95% CI: -3.7 to -0.3; p=0.021). Degradation was significant at multiple pool sizes (k = 15-50; p<0.05). This difference between architectures demonstrates that optimization strategies derived from one architecture may actively harm another.

## Discussion

Our findings reveal a fundamental disconnect between benchmark performance and clinical reasoning capability in medical large language models (LLMs). LLMs achieving 85-90% accuracy on medical licensing examinations exhibited dramatic instability when processing realistic oncology cases.^3,16^ OpenBioLLM tumor staging accuracy shifted from 45.9% to 99.1% based solely on prompt format, MedGemma generated a definitive staging 100% of the time despite intentionally insufficient clinical information, and GPT-5 was only 7% accurate on understanding clinical tumor stage IVB (**Figure 1c**). Concordance with AJCC 8/9^th^ edition guidelines reached only 26.8%-31.0% for open-source models versus 97.5% for GPT-5 (**Figure 2b**), and MedGemma and OpenBioLLM are dependent on correct staging for correct treatment response (**Figure 2d-e**). These findings align with growing evidence questioning standard evaluation paradigms. Script concordance testing reveals that models achieving near-expert scores on multiple-choice examinations consistently underperform clinicians on tasks requiring probabilistic reasoning under uncertainty, with systematic overconfidence manifesting as bias toward extreme responses.^19^ These limitations have prompted calls to retire medical licensing examinations as primary benchmarks, given their fundamental inadequacy as metrics for clinical utility.^5^ Evaluations using real patient data from MIMIC-IV demonstrate a 16-25% gap between LLM and physician diagnostic accuracy when models must gather clinical information rather than select from provided options.^5^ The MedS-Bench evaluation spanning 11 clinical tasks found that even GPT-4 struggles with complex reasoning despite strong performance on medical question-answering, underscoring the gap between benchmark success and clinical demands.^20^ Our perturbation analysis extends these observations by demonstrating that instability emerges not only across task types but within single clinical scenarios when superficial input characteristics change.

Our sparse autoencoder analysis provides a representational explanation for these phenomena. MedGemma organizes polysemous medical terms hierarchically, with the top-10 features per token showing 77.8% overlap regardless of clinical context, encoding word identity, while low-magnitude features showed only 16.9% overlap, encoding contextual meaning (**Figure 4a**). OpenBioLLM distributed information uniformly, with 10–18% overlap at all feature magnitudes. This architectural difference (hierarchical versus distributed) means that when a clinical decision support system processes a query about “arrest,” MedGemma’s high-magnitude features activate similarly for respiratory arrest and cardiac arrest, potentially retrieving cardiac protocols when respiratory guidance is needed. Ablation experiments confirmed these context-encoding features are causally necessary, degrading next token prediction by 16.4 nats in MedGemma and 6.7 nats in OpenBioLLM (**Figure 4b**). These effects were domain-general, in which degradation in radiological contexts was statistically similar to cardiac domains (**Figure 4d**), suggesting both architectures rely on distributed feature coalitions rather than localized “medical knowledge” detectors (**Figure 4e**). Medical knowledge therefore is shown to be compartmentalized across thousands of specialized features, with dental terminology in one feature, neurological gait in another (Table 1), reinforcing theoretical models where neural networks pack more features than neurons.^21^ This organization therefore challenges safety approaches that rely on detecting or steering broad content categories. The practical consequences of this representational heterogeneity are significant. Retrieval-augmented generation has emerged as a leading approach for grounding LLM outputs in verified clinical knowledge, with studies demonstrating that RAG can improve accuracy to 96.4% with zero hallucinations in surgical fitness assessment, significantly outperforming human clinicians.^22^ However, RAG effectiveness varies substantially across LLM architectures, and systematic reviews identify a lack of standardized evaluation frameworks for RAG-based clinical applications.^23,24^ Additionally, RAG cannot reliably alter LLM behavior.^8,9^ Our two-stage retrieval intervention, designed to filter identity-encoding features during re-ranking, improved MedGemma’s precision by 10.2% (p=0.022, paired t-test) but degraded OpenBioLLM’s performance by 2.0% (p=0.021; **Figure 4f**). This 12.2% difference demonstrates that optimization strategies derived from one architecture may actively harm another, consistent with evidence that prompt engineering effectiveness varies dramatically across LLM families (*κ*= - 0.002 to0.984)^25^ and that safety fine-tuning produces architecture-dependent outcomes.^26^ The mechanistic basis for why these architectures developed different encoding strategies remains unclear. Possible factors include training data composition, optimization dynamics, or model capacity.^27–30^ However, regardless of origin, our findings demonstrate that these representational differences have direct, measurable consequences on clinical AI safety. Current regulatory frameworks and institutional validation protocols typically assume model-agnostic safety properties^31^, but our results suggest this assumption is incorrect.

Due to the significant storage and computational demands of this work, we analyzed two model architectures. Additional architectures (Mistral, Phi, Qwen, GPT-OSS) may exhibit distinct patterns. Our SAE analysis used 64-token context windows, following prior work,^11^ which may potentially underrepresent document-level semantic relationships. MIMIC-IV represents single-center intensive care documentation from Beth Israel Deaconess Medical Center, limiting generalization to outpatient or international settings. The three synthetic oncology cases and retrieval tasks, while demonstrating mechanistic principles, represent simplified proxies for production clinical decision support systems.

In conclusion, our results demonstrate that benchmark equivalence does not imply functional equivalence, interpretability findings do not transfer across architectures, and safety interventions must be validated architecture-by-architecture, generalizable to other high-stakes domains (e.g., scientific research). Current regulatory frameworks and institutional validation protocols typically assume model-agnostic safety properties. Our findings indicate this assumption requires reconsideration. We recommend that healthcare institutions evaluating medical AI systems should test clinical reasoning stability under realistic perturbations, validate retrieval and safety interventions on the specific architecture being deployed, and incorporate mechanistic interpretability methods to characterize how candidate systems encode clinical knowledge.

## Methods

### Synthetic oncology case evaluation

To assess clinical reasoning stability under controlled conditions, we developed three synthetic oncology cases in collaboration with two board-certified physicians: a urethral carcinoma case (UC), a non-small cell lung cancer follow-up case (NSCLC-FU), and a non-small cell lung cancer initiate treatment case (NSCLC-IT). Cases were constructed in SOAP (Subjective, Objective, Assessment, and Plan) format, with the Subjective and Objective (S&O) sections serving as LLM input, with Assessment and Plan (A&P) sections serving as expected LLM outputs. To understand the baseline variability in LLMs, we measure similarity between the embeddings of the LLM generated A&P and the physician A&P. We use BioLORD-2023^32^ to generate the embeddings from the notes. For each case, we systematically removed individual medical terms (Table S1) from the input S&O prompt, generating 111 perturbations for Urethral Carcinoma (UC), 142 for non-small cell lung cancer initiate treatment (NSCLC-IT), and 102 for non-small cell lung cancer follow-up (NSCLC-FU). Then, we measured (i) cosine similarity when no terms are removed between LLM-generated A&P embeddings versus physician-written A&P reference embeddings, (ii) tumor staging accuracy, and (iii) treatment recommendation categories (management with or without perioperative management, systemic therapy, radiation with or without chemotherapy, further workup needed, or ambiguous) per NCCN treatment guidelines. We tested three prompting strategies: full A&P generation, direct staging/treatment queries, and chain-of-thought from TNM staging to clinical staging to treatment recommendation. For chain-of-thought, we also measured concordance between model-generated TNM staging and the model-generated clinical staging versus AJCC 9^th^ edition-(or 8^th^ edition for UC)-clinical staging. Detailed experiments were conducted on GPT-5, MedGemma-27B-Text-IT, and OpenBioLLM-Llama3-70B to assess consistency and accuracy across model families.

### Sparse autoencoder study design and data sources

We conducted a computational study applying top-k SAE analysis to medical LLMs using deidentified clinical notes from the MIMIC-IV-Note (Medical Information Mart for Intensive Care, version 2.2) database. MIMIC-IV-Note is a publicly accessible critical care database containing free-text clinical notes from Beth Israel Deaconess Medical Center from 2008 to 2019, comprising 331,794 deidentified discharge summaries about chief complaint, history of present illness, hospital course, physical exam findings, and discharge diagnoses across 145,915 unique patients.^17^ We also utilized the MIMIC-IV-Ext-BHC extension database, which provides 270,033 clinical notes due to standardized Brief Hospital Course sections derived from discharge summaries suitable for language model analysis.^18^ From this corpus, we randomly sampled 50,000 discharge notes for SAE training, ensuring representation across the full temporal range and clinical diversity of the database. This study was conducted under the MIMIC-IV data use agreement and did not require additional institutional review board approval, as all data were previously deidentified.

### Medical large language model selection for sparse autoencoder study

We selected two leading medical LLMs for analysis based on benchmark performance, open-source accessibility, and representativeness of current medical AI systems.^3,16^ MedGemma-27B-Text-IT is a 27-billion parameter instruction-tuned model developed by Google Research, built on the Gemma 3 architecture. It utilizes a 128,000-token context length supported by an interleaved local-global attention mechanism and grouped-query attention.^3^ The model was fine-tuned on medical text and question-answer pairs, achieving 87.7% accuracy on MedQA (zero-shot) and state-of-the-art performance across multiple medical benchmarks.^3^ OpenBioLLM-Llama3-70B was developed by Saama AI Labs, fine-tuned from Meta’s Llama-3-70B-Instruct using Direct Preference Optimization (DPO) and a two-phase fine-tuning process on a curated dataset of over 3,000 healthcare topics.^4^ At the time of analysis, this model ranked among the top performers on the Open Medical-LLM Leaderboard, achieving 86.06% average accuracy across medical subsets of the Massive Multitask Language Understanding benchmark.^16^ Together, these models represent distinct architectural approaches (native medical pretraining versus domain-specific fine-tuning) and parameter scales, enabling assessment of whether observed representational patterns generalize across medical language model designs.

### Activation extraction

For each clinical note, we tokenized the text using the respective LLM’s tokenizer and performed a forward pass through the LLM, extracting intermediate layer activations at each token position. We extracted activations from layer 52 of MedGemma-27B (of 61 total layers including layer 0) and layer 67 of OpenBioLLM-70B (of 79 total layers including layer 0).

Activations were extracted using context windows of 64 tokens with a stride of 64 in bfloat16 precision. Activations were L2-normalized before storage. We preserved character-level position offsets and document identifiers for each token to enable downstream alignment with named entity annotations. We used a 95% / 5% split for training and validation.

### Sparse autoencoder architecture and training

SAEs learn dictionaries of features by reconstructing dense LLM activation vectors (see supplementary methods) using sparse combinations of learned latent directions. We employed the Top-K SAE, which enforces sparsity by retaining only the K-highest magnitude latent activations during the forward pass rather than relying on L1 regularization, following the training scheme of OpenAI.^11^ We use a 12x expansion factor when training our SAE (65,536 latent for MedGemma, 98,304 latent for OpenBioLLM), with a fixed sparsity *k* of 128. These expansion factors and sparsity levels were selected based on prior work demonstrating that 10-15x expansion with approximately 100 active features yields interpretable decompositions with acceptable reconstruction fidelity^11^ and our own small-scale experiments (**Figures S1-4**; Supplementary Methods). The SAE was trained using the Adafactor optimizer with a learning rate 2*10^-4^, a micro-batch of 32,768 tokens, gradient clipping at 1.0, and an auxiliary loss coefficient of 0.03125 with auxiliary sparsity of 512 for dead latent recovery (**Table S1**). Training proceeded for 200,000 steps with validation every 2,000 steps. We train the SAE for approximately 1 billion tokens. We report results at this convergence point.

### Feature normalization

For fair feature comparisons, we scale the SAE encoder and decoder weights by the maximum activation value in a scan of 100,000 tokens from MIMIV-IV-Note for each feature.

### Named entity recognition and sense disambiguation

To ground SAE features in clinical semantics, we annotated all clinical notes with named entity labels using a fine-tuned named entity recognition model, gliner-biomed-large-v1.0 NER.^33^ We classified spans into medical entity categories: anatomical structure, bodily fluid or secretion, cardiovascular disorder, clinical finding or symptom, dermatological lesion, gastrointestinal disorder, laboratory measurement, medical procedure or intervention, medication or therapeutic agent, musculoskeletal disorder, neurological or mental disorder, radiological finding, renal or kidney disorder, respiratory or pulmonary disorder, and administrative healthcare event. Entity annotations were aligned to SAE activations using character-level offsets, matching each token to overlapping named entity spans. Tokens outside medical entity spans were labeled as general text. For medical terms with multiple distinct meanings (“polysemous”), we developed a sense disambiguation procedure mapping named entity labels to semantic categories. We identified 12 high frequency polysemous and 4 monosemantic medical terms in our dataset (**Tables S3-4**). Tokens for which the sense could not be resolved from the named entity context were removed from the analysis.

### Feature decomposition and polysemantic analysis

We measured the unique latent patterns for each token to determine how the medical LLMs represent polysemous medical terms versus monosemantic medical terms (see supplementary methods for annotation strategy). We compute the pairwise Jaccard similarity between feature sets activated for tokens in different sense categories (cardiac arrest versus respiratory arrest). High overlap indicates features that activate regardless of sense (i.e., lexical representation), and low overlap indicates sense-discriminating features (i.e., semantic representation). We fit univariate L2-regularized logistic regression probes for each latent using Adam optimization to distinguish medical entities from background text. For each latent, we computed the area under the receiver operating characteristic curve (AUROC) for binary classification of each entity type. Latent for each sense of each polysemous term were also fitted using a generalized linear mixed model to measure the entropy of each polysemous term.

### Ablation and causal analysis

To assess whether identified features causally influence LLM predictions, we performed activation ablation experiments measuring the change in cross-entropy loss (Δ*CE*) when specific latent directions were zeroed:

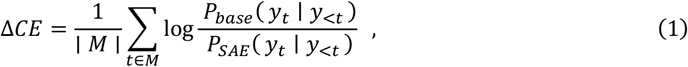

where M represents the valid tokens. Positive ΔCE indicates prediction degradation from the intervention. For each medical entity category, we identified sense-linked latent from probe analysis and measured Δ*CE* when ablating those features during forward passes on held-out clinical notes. We modeled the ablation effects using mixed-effects regression with random intercepts for document and fixed effects for medical context category, quantifying which clinical domains exhibited feature-mediated prediction changes.

### Retrieval experiments for sense disambiguation

To validate the functional relevance of lexical features versus semantic features, we designed a two-tier structure where we re-rank high magnitude latent (lexical identity) and lower magnitude latent (contextual semantics). Given a query token such as “discharge”, we retrieve the nearest-neighbor tokens of the same word and evaluate whether the retrieved tokens shared the query’s semantic sense. Terms were only included for analysis if they had ≥30 total token instances and ≥2 senses with ≥5 instances each. For each query token, we computed cosine similarity against all-other same word tokens using three feature subsets: full (all 128 latent), lexical only (top-10 latent), and semantic-only (latent 11-128). The two-stage re-ranking retrieves the top-k candidates by full similarity first, then re-ranks these *k* candidates by semantic-only similarity. We swept candidate pool sizes *k* ∈ {5,10,15,20, …,100} to identify optimal retrieval depth. Performance was measured by precision at rank 1 (P@1) and mean average precision (MAP). For each *k*, we computed the improvement over baseline (Δ*P*@1 = *P*@1_re−rank_ − *P*@1_full similarity_) across all query tokens. Statistical significance was assessed using paired t-tests comparing re-ranked versus baseline P@1 for each query, with 95% confidence intervals computed as 1.96*SE.

## Data Availability

The MIMIC datasets used for our work can be found at physionet.org/content/mimic-iv-note/2.2/ and physionet.org/content/labelled-notes-hospital-course/1.2.0/.Any intermediate files can be produced by the corresponding authors upon reasonable request and signing of any necessary data use agreements.

## Acknowledgements

This work was funded by the National Institutes of Health (NIH; T32GM139784, U54AG075931) and by the Pelotonia Institute of Immuno-Oncology. We thank Dr. Fei He for his feedback regarding the manuscript.

