## Supplemental for "Understanding Clinical Reasoning Variability in Medical Large Language Models: A Mechanistic Interpretability Study"

***Supplementary Appendix***

Mirage Modi[1], Jordan E. Krull[1,2,$], Donte Johnson[1], Xiaoying Wang[1,2], Timothy D. Gauntner[2,3], Mingjia Li[3], Hao Cheng[1], Anjun Ma[1,2], Ping Zhang[1,4], Daniel G. Stover[3], Zihai Li[2,3], Qin Ma[1,2,$]

[1]Department of Biomedical Informatics, The Ohio State University College of Medicine, Columbus, OH, USA
[2]Pelotonia Institute for Immuno-Oncology, The Ohio State University Comprehensive Cancer Center – James Cancer Hospital and Solove Research Institute, Columbus, OH, USA
[3]Division of Medical Oncology, Department of Internal Medicine, The Ohio State University Comprehensive Cancer Center – James Cancer Hospital and Solove Research Institute, Columbus, OH, USA
[4]Department of Computer Science and Engineering, The Ohio State University, Columbus, OH, USA

**Urethral Carcinoma**

Subjective: Patricia Gateau is a 61-year-old female with PMHx significant for HTN, HLD, CAD s/p PCI with stent to LAD, mild sensorineural hearing loss who presents to establish care with GU Med Onc for newly diagnosed urethral carcinoma. She had initially presented to her PCP with vaginal bleeding, an enlarging vaginal mass and several weeks of urinary frequency. Today she reports that vaginal bleeding has stopped, however, she still has urinary frequency and now notes a weak urine stream.

Objective: Vitals signs: weight 91.1 kg, height 175 cm, BP 152/74. HR 76, RR 18, O2 saturation 95% on RA

Physical exam: General: alert, no acute distress, ECOG 1; Heart: RRR, normal S1/S2, no murmurs; Lungs: CTA b/l; Abd: Soft, nontender, non-distended; GU: On bi-manual exam there is a firm, fixed periurethral mass that appears to invade the anterior vagina; MSK: No LE edema; Skin: no rash; Neuro: No gross neurological deficits, CN 2-12 intact.

Labs significant for normal creatinine at 0.80 mg/dL.

CT urogram demonstrates a distal urethral lesion measuring 1.6 x 1.2 x 2.3 cm, but no evidence of pelvic lymphadenopathy or metastatic disease.

Pathology from core biopsy of periurethral mass reveals an invasive poorly differentiated carcinoma with squamous differentiation. IHC stains are positive for CK7, p63, p16, p40 and GATA3 and negative for CK20.

A&P: 61 y.o. female with hx of HTN, HLD, CAD s/p PCI with stent to LAD, mild sensorineural hearing loss and newly diagnosed p16+ poorly-differentiated urethral carcinoma with squamous differentiation. This appears to be a clinical stage III (T3N0M0) urethral carcinoma. We discussed the role of chemotherapy in definitive management of stage III urethral carcinoma, either as neoadjuvant chemotherapy followed by surgical resection of her mass or as concurrent chemoradiotherapy. Concurrent chemoradiotherapy with a radiosensitizing chemo regimen (either single agent cisplatin, low dose gemcitabine or 5-FU + mitomycin) would be the preferred approach, except in urothelial cell carcinoma, where neoadjuvant chemotherapy (with either gemcitabine + cisplatin or dose-dense MVAC) followed by consolidative surgery would be recommended. Since her pathology shows p16+ poorly differentiated carcinoma with squamous differentiation (and NOT urothelial cell carcinoma) I would favor concurrent chemoradiation. Given her hx of sensorineural hearing loss it will be important to obtain baseline audiometry to assess for cisplatin eligibility. Her dysuria/urinary frequency likely represents urinary tract outlet obstruction due to urethral mass vs UTI vs overactive bladder vs interstitial cystitis.

- Referral to Urologic Oncology
- Referral to Radiation Oncology
- Referral to Audiology (to assess for cisplatin eligibility)
- Bladder scan/PVR in clinic today (to assess for urinary retention / outlet obstruction that may necessitate temporary placement of Foley catheter)
- Return to GU Med Onc clinic in 4 weeks for follow up and to review recommendations from Urology and Rad Onc

**Non-small cell lung cancer follow-up**

Subjective:
This a 58 y.o. male with discoid lupus, extensive smoking history (40 pack-years, quit 2019), and newly diagnosed squamous cell carcinoma of the lung is here for further management. He was initially hospitalized for hemoptysis on 12/21/23 locally. CT on 12/23/23 showing LLL completely atelectatic, small left pleural effusion, mildly enlarged AP node, small bilateral adrenal lesions (1.5 cm on the right with hounsfield of 7, and 1.3 cm on the left with Hounsfield 20) concern for adenoma. 12/29/23: Bronch, distal left main bronchus, just prior to left upper lobe, a fungating bleeding mass was visualized, with 60-70% obstruction to the airway. This lesion was biopsied. Per report, no abnormal LN station were seen. Biopsy positive for squamous cell carcinoma. He also had an arterial embolization done on 12/26/23.
Interval History
Patient is here for his initial follow at OSU.
Overall he is feeling relatively well. Since the initial hospitalization at the end of December, he has not had further hemoptysis. Denies any significant chest pain. Has some intermittent mild cough. Overall, he is feeling stable.
He has a history of discoid lupus involving the face and chest chest skin, on plaquenil for 6 months back in 1980s.
He denies any headache, vision changes, hearing changes, dysphagia, nausea, vomiting, abd pain, constipation, diarrhea, dysuria, focal weakness, leg swelling, peripheral neuropathy, fever, chill.

Objective:
**Vital Signs:** BP 160/73 (BP Location: Right arm, BP Position: Sitting) | Pulse 78 | Temp 97.7 °F (36.5 °C) (Oral) | Resp 14 | Ht 1.829 m (6') | Wt 127 kg (280 lb) Comment: with shoes and jacket on | SpO2 95% | BMI 37.97 kg/m² | Smoking Status Former
**ECOG performance status: 0**
**Constitutional:** male alert and oriented in no acute distress.
**Eyes:** No scleral icterus. EOMI.
**ENMT:** No oropharyngeal lesions or thrush.
**Neck:** no masses, symmetrical.
**Respiratory:** Normal effort. Clear to auscultation: A&P bilaterally, no crackles/rhonchi/wheezes
**Cardiovascular:** Normal S1/S2, regular rate and rhythm, no murmurs, gallops, rubs. No peripheral edema bilaterally
**Abdomen:** soft, non-tender. Bowel sounds normal.
**Musculoskeletal:** no digital cyanosis or clubbing.
**Skin:** No rashes or lesions.
**Lymph nodes:** Cervical and supraclavicular nodes normal.
**Neurologic:** Cranial nerves grossly intact. No focal deficits.
**Psychosocial:** Affect appropriate for situation. Alert and oriented to person, place and time.

12/29/23: Bronch with left main bronchus mass biopsy positive for squamous cell carcinoma.
**CT Chest:**
**12/23/23:**
1. The left lower lobe is completely atelectatic and there is abrupt
termination of the proximal left lower lobe bronchus concern for obstructive from a mass.
2. mildly enlarged mediastinal lymph node.
Small bilateral adrenal lesion.

**CT Abdomen:**
**None**

**MRI Brain:**
**None**

**PET/CT:**
None

A&P:
This is a pleasant 58 y.o. male with discoid lupus, extensive smoking history, and newly diagnosed squamous cell carcinoma involving the left main bronchus.

**Abbreviated Plan Summary**

- **Stage:** Staging is incomplete
- **Primary disease site**: Left Lower lobe
- **Histology:** Squamous Cell carcinoma
- **Significant Molecular Status:** Unknown
- **PD-L1%:** Unknown
- **Prior cancer therapies:** None
- **Last staging (imaging):** 12/23/23, incomplete
- **Current Therapy:** None
- **Anticipate next treatment options: TBD**

1. Stat MRI of brain, CT chest/abd/pelvis, and PET scan to complete staging.
2. Guardant 360 from peripheral blood today. Additionally, I will add molecular testing, including PD-L1, to his LLL lung mass biopsy that was done on 12/29/23.
3. We have discussed the importance and time frame of completing the remaining staging and molecular workup.
4. If no metastatic disease is identified, will refer him to thoracic surgery for surgical evaluation.
5. RTC in 2 weeks

Discoid lupus erythematosus
-involving the skin on the face and chest
-per patient, only required a brief 6 months of treatment back in 1980s with plaquenil. Has not needed any additional treatment since.
-will reach out to dermatology if immunotherapy is needed.

**Non-small cell lung cancer initiate treatment**

Subjective:
This is a 60 y.o. male, former heavy smoker, with newly diagnosed metastatic carcinoma, favoring squamous cell carcinoma from pulmonary origin is here for further management. He initially presented in the ED on 4/24/25 for severe left flank pain and was found to have spontaneous subcapsular renal hematoma. CT on 4/24/25 significant for moderate sized left subcapsular hematoma measuring 2.5 cm, and right renal cortical cysts measuring 2.5 cm; additionally, there is a 1.7 cm probable liver cyst, and a right 1.6 cm adrenal nodule. Follow up MRI on 4/30/25 showing exophytic simple cyst in the right kidney, and heterogenous enhancing

lesion in the left upper pole of left kidney with hematoma; enhancing 2.3 cm lesion in the liver. CT chest on 5/5/25 showing right upper lobe mass measuring 4.7 cm, mediastinal and hilar lymphadenopathy with scattered pulmonary nodules. A liver biopsy was done on 5/7/25, showing poorly differentiated carcinoma (PD-L1 99% in house, and 60% from Tempus; CPS 60 from tempus, +TP53, NOTCH2, and NF2). He presented in the ED again for severe right flank pain, and was found to have bilateral subcapsular hematoma in both kidney, with AKI that required brief dialysis.

PET scan as part of workup for carcinoma of unknown primary on 5/22/25 showing significantly increased uptake in the left tonsils, left lung mass and thoracic lymph nodes, liver. EGD and colonoscopy was done, few polyps (tubular adenomas and hyperplastic polyps) were seen and negative for malignancy.

A CT guided lung biopsy was done on 5/25/25 for the right upper lobe lung mass. Positive for non-small cell carcinoma, favoring squamous cell carcinoma (TPS 60%, tumor morphology is similar to that liver biopsy). Tempus tissue origin was run from the liver sample, 46% probability being pancreatic cancer and 11% pulmonary squamous cell carcinoma, and 11% probability from head and neck.

Interval History
Patient is here with his wife today for follow up.

Overall, he is feeling better since discharged from the hospital. He is able to ambulate without difficulty. Pulmonary symptoms is stable, without significant worsening of chest pain, dyspnea, hemoptysis.

He still has difficulty with urinary and still has indwelling foley catheter.

He denies any headache, vision changes, hearing changes, dysphagia, nausea, vomiting, abd pain, constipation, diarrhea, dysuria, focal weakness, leg swelling, peripheral neuropathy, fever, chill.

Objective:
Vital Signs: BP 130/78 (BP Location: Left arm, BP Position: Sitting) | Pulse 70 | Temp 98.2 °F (36.8 °C) (Oral) | Resp 18 | Ht 1.803 m (5' 11") Comment: reported by pt | Wt 100.2 kg (221 lb) Comment: with shoes on | SpO2 98% | BMI 30.82 kg/m² | Smoking Status Heavy Smoker
ECOG performance status: 1
Constitutional: male alert and oriented in no acute distress.
Eyes: No scleral icterus. EOMI.
ENMT: No oropharyngeal lesions or thrush.
Neck: no masses, symmetrical.
Respiratory: Normal effort. Clear to auscultation: A&P bilaterally, no crackles/rhonchi/wheezes
Cardiovascular: Normal S1/S2, regular rate and rhythm, no murmurs, gallops, rubs. No peripheral edema bilaterally
Abdomen: soft, non-tender. Bowel sounds normal.
Musculoskeletal: no digital cyanosis or clubbing.
Skin: No rashes or lesions.
Lymph nodes: Cervical and supraclavicular nodes normal.
Neurologic: Cranial nerves grossly intact. No focal deficits.
Psychosocial: Affect appropriate for situation. Alert and oriented to person, place and time.

**Pathology:**
5/25/25: CT guided biopsy of the lung mass
**Non-small cell carcinoma, favor squamous cell carcinoma**

5/7/25:
Pathologic Diagnosis A. Liver, lesion, biopsy: · Poorly differentiated carcinoma

**CT chest**
**5/5/25**
**4.3 CM right upper lobe mass. Scattered pulmonary nodules. Enlarged mediastinal lymph nodes.**

**CT abd pelvis**
5/23/25:
Known progressing pulmonary metastases. Stable subcapsular renal hematoma bilaterally.

**PET/CT**
5/22/25
Significantly increased uptake in the left tonsils, left lung mass and thoracic lymph nodes, liver.

**Brain MRI**
5/5/25
No metastatic disease

A&P:
This is a pleasant 60 y.o. male, former heavy smoker, who presents today for evaluation of metastatic carcinoma, favoring squamous cell carcinoma. Base on the image finding with large right upper lobe mass with mediastinal lymphadenopathy, presumed from the lung.

**Abbreviated Plan Summary**
- **Stage:** Stage IV
- **Primary disease site**: Presumed right upper lung
- **Histology:** Poorly differentiated carcinoma, favoring squamous cell
- **Significant Molecular Status:** TP53, NF2, CDKN2A, **HER2 IHC +3**
- **PD-L1%:** TPS 60, CPS 60
- **Prior cancer therapies:** None
- **Last staging (imaging):** 5/22/25
- **Current Therapy:** None
- **Anticipate next treatment options:** Pembrolizumab monotherapy

1. Based on the image finding with large right upper lobe mass with mediastinal lymphadenopathy, presumed from the lung. However, PET also showing some uptake in the tonsillar with cervical lymphadenopathy, can not completely rule out of Head and Neck origin as well. Tempus tissue origin showing equal 11% probability of being from lung and head/neck. We have discussed in lengthy detail regarding the origin of the cancer, and based on the image finding, overall favoring NSCLC.
2. I have reviewed his PET on 5/22/25, and there is visible growth in many lesion just comparing to few weeks ago. His tumor showing high expression of TPS, and I recommend pembrolizumab monotherapy. Especially given multiple hospitalizations, overall performance

status, spontaneous bilateral subcapsular hematoma; the risk of chemo, especially increased bleeding risk of his subcapsular hematoma in light of cytopenia can be high.
3. In the Keynote 024 study, patients with a PD-L1 TPS of 50% or higher had a median overall survival (OS) of 26.3 months compared to 13.4 months with chemotherapy (HR 0.62, 95% CI 0.48–0.81). The 5-year OS rate was 31.9% for pembrolizumab versus 16.3% for chemotherapy. In terms of objective response, 4.5% had CR, 41.6% had PR, and 24% had stable disease. Median PFS was 7.7 months for pembrolizumab. Grade 3 or higher immune-related adverse events (irAEs) occurred in 13.6% of patients treated with pembrolizumab (PMID: 33872070). In addition, pembrolizumab will also cover for metastatic squamous cell carcinoma from head and neck with high CPS.
4. Due to concern for long commute, he is scheduled to see Dr K locally next week for treatment.

Spontaneous subcapsular hematoma of both kidneys:
-resulting in AKI that briefly needing dialysis in May 2025
-I have discussed the case with Dr G from GU oncology. Overall, low concern for RCC currently. However, will benefit from tissue sampling in the future if feasible
-will have him follow up with GU oncology in the clinic.

Urinary obstruction:
-follow up with urology locally
-has indwelling foley

**System Prompt**

"""

You are a leading expert physician in Medical Oncology. Your goal is to formulate an Asessment & Plan (A&P), with the exact clinical stage diagnosis in TNM staging and the best definitive management for your patient's visit, using the provided Subjective and Objective note from your patient encounter.

Please keep in mind the patient encounter note may be missing pieces of information. If the patient encounter note contains [MISSING INFO], then you can assume that information is missing. For phrases where that is not present, missing information may or may not be present. Regardless, you are still responsible for fully understanding the note. Your patient encounter note (Subjective and Objective) is summarized below:

[S&O]

You need to be very specific in your response, providing your rationale and using medical terminology where appropriate.

"""

**User Prompt**

*Direct*

“please only tell me the best definitive management option for the patient if applicable. Write ‘unknown’ anywhere where the answer is not known.”

“please only tell me the exact TNM clinical staging for the patient. Write ‘unknown’ anywhere where the answer is not known.”

“Would you like to perform any additional tests? Only suggest additional tests if you think it is strictly necessary in significantly informing your definitive management plan. Write ‘unknown’ anywhere where the answer is not known.”

*Full A&P (example from NSCLC-FU case)*

"""

Based on your goal and this note,

1. Provide a detailed clinical assessment

2. Provide a detailed clinical plan based on your assessment

3. Identify the best option for definite management if applicable.

4. Would you like to perform any additional tests? Only suggest additional tests if you think it is strictly necessary in significantly informing your plan.

Draft your response using the A&P style format note I have provided below. Strictly fill out and adhere to the instructions provided in the areas enclosed in “[]”.

NOTE TO FILL OUT:

A&P:

[Patient summary of presentation].

Abbreviated Plan Summary

[Abbreviated Plan Summary, write "unknown" anywhere where the answer is not known.]

- Stage: [Stage, If this cannot be determined from the provided information, write "unknown". If you think that the staging is "unknown", write "TBD" if it is pending completion. Otherwise, strictly write the exact TNM staging.]
- Primary disease site: [Primary disease site]
- Histology: [Histology diagnosis]
- Significant Molecular Status: [Significant Molecular Status]
- PD-L1%: [PD-L1%]
- Prior cancer therapies: [Prior cancer therapies]
- Last staging (imaging): [Last staging (imaging) date and status]
- Current Therapy: [Current therapy]
- Anticipate next treatment options: [Anticipate next treatment options. If to be decided, write TBD.]

1. [Additional imaging only if strictly necessary].

2. [Additional testing only if strictly necessary].

3. [What you would like to discuss with the patient].

4. If no metastatic disease is identified, will [plan if no metastatic disease found].

5. [return to clinic timeframe in weeks].

[The other disease of the patient]

-[description of disease manifestation on patient]

-[tried treatments by patient].

-[referral plan].

"""

**Chain of Thought**

*Prompt (Urethral Carcinoma Example)*

"""

You are a leading expert physician in Medical Oncology. Your goal is to formulate an Asessment & Plan (A&P), with the exact clinical stage diagnosis in TNM staging and the best definitive management for your patient's visit, using the provided Subjective and Objective note from your patient encounter. Please keep in mind the patient encounter note may be missing pieces of information. If the patient encounter note contains [MISSING INFO], then you can assume that information is missing. For phrases where that is not present, missing information may or may not be present. Regardless, you are still responsible for fully understanding the note. Your patient encounter note (Subjective and Objective) is summarized below:

[S&O]

You need to be very specific in your response, providing your rationale and using medical terminology where appropriate.

You are a clinical decision-support assistant.

You may use a private scratchpad to reason, but NEVER reveal your notes.

Only output a single JSON object that strictly matches this JSON Schema:

{"type": "object", "properties": {"guidelines_used": {"type": "array", "items": {"type": "string"}}, "tnm": {"type": "string"}, "clinical_stage": {"type": "string"}, "definitive_management": {"type": "string"}, "brief_justification": {"type": "string"}}, "required": ["guidelines_used", "tnm", "clinical_stage", "definitive_management", "brief_justification"], "additionalProperties": false}

Rules:

- If TNM cannot be determined from the provided info, set "tnm": "unknown".

- If you believe staging is pending completion, use "TBD"; otherwise use the exact TNM string.

- "clinical_stage" must reflect the TNM (e.g., "Stage IIIB").

- "brief_justification" is ≤ 2 sentences. Do NOT include step-by-step reasoning.

- List the guideline titles/versions in "guidelines_used" (e.g., "AJCC 8th ed.").

- Output JSON only. No prose or markdown.

Fill the JSON fields by addressing, in order:

1) Which guidelines you used to determine staging and management.

2) Exact TNM (or "unknown"/"TBD" per the rules).

3) Resulting clinical stage.

4) Best definitive management option based on the above.

Return JSON only.

"""

**Sampling Strategy**

We employed self-consistency decoding to improve response reliability. Rather than relying on a single greedy or low-temperature generation, we sampled $n = 5$ independent responses using nucleus sampling with temperature $\tau = 0.6$ and top-$p = 0.9$. We performed majority voting on the definitive_management field (the primary clinical decision of interest). The complete response corresponding to the majority answer was returned as the final output. This approach leverages the observation that correct reasoning paths tend to converge on consistent answers, while errors are more randomly distributed across samples.[1]

### SUPPLEMENTARY TABLES

Table S1. SOAP terms removed from case

| Case | Term Removed |
| --- | --- |
| Urethral Carcinoma | BASELINE (*No removals)* |
| Urethral Carcinoma | PMHx |
| Urethral Carcinoma | urethral carcinoma |
| Urethral Carcinoma | HTN |
| Urethral Carcinoma | HLD |
| Urethral Carcinoma | CAD s/p PCI with stent to LAD |
| Urethral Carcinoma | CAD |
| Urethral Carcinoma | PCI |
| Urethral Carcinoma | stent |
| Urethral Carcinoma | LAD |
| Urethral Carcinoma | mild |
| Urethral Carcinoma | sensorineural hearing loss |
| Urethral Carcinoma | sensorineural |
| Urethral Carcinoma | hearing |
| Urethral Carcinoma | loss |
| Urethral Carcinoma | hearing loss |
| Urethral Carcinoma | establish care with GU Med Onc |
| Urethral Carcinoma | newly diagnosed urethral carcinoma |
| Urethral Carcinoma | enlarging |
| Urethral Carcinoma | vaginal bleeding |
| Urethral Carcinoma | enlarging vaginal mass |
| Urethral Carcinoma | vaginal mass |

| | |
|---|---|
| Urethral Carcinoma | mass |
| Urethral Carcinoma | bleeding |
| Urethral Carcinoma | vaginal |
| Urethral Carcinoma | urinary frequency |
| Urethral Carcinoma | urinary |
| Urethral Carcinoma | frequency |
| Urethral Carcinoma | several weeks of urinary frequency |
| Urethral Carcinoma | several weeks |
| Urethral Carcinoma | vaginal bleeding has stopped |
| Urethral Carcinoma | stopped |
| Urethral Carcinoma | weak |
| Urethral Carcinoma | weak urine stream |
| Urethral Carcinoma | urine stream |
| Urethral Carcinoma | 91.1 kg |
| Urethral Carcinoma | 175 cm |
| Urethral Carcinoma | 152/74 |
| Urethral Carcinoma | 76 |
| Urethral Carcinoma | 18 |
| Urethral Carcinoma | 95% on RA |
| Urethral Carcinoma | alert, no acute distress |
| Urethral Carcinoma | alert |
| Urethral Carcinoma | no acute distress |
| Urethral Carcinoma | ECOG 1 |
| Urethral Carcinoma | RRR, normal S1/S2, no murmurs |
| Urethral Carcinoma | RRR |
| Urethral Carcinoma | normal S1/S2 |
| Urethral Carcinoma | no murmurs |
| Urethral Carcinoma | Soft |
| Urethral Carcinoma | nontender |
| Urethral Carcinoma | non-distended |
| Urethral Carcinoma | bi-manual exam |
| Urethral Carcinoma | firm |
| Urethral Carcinoma | fixed |
| Urethral Carcinoma | periurethral |
| Urethral Carcinoma | invade the anterior vagina |
| Urethral Carcinoma | anterior |
| Urethral Carcinoma | vagina |
| Urethral Carcinoma | CTA b/l |
| Urethral Carcinoma | Soft, nontender, non-distended |

| | |
|---|---|
| Urethral Carcinoma | On bi-manual exam there is a firm, fixed periurethral mass that appears to invade the anterior vagina |
| Urethral Carcinoma | neurological deficits |
| Urethral Carcinoma | deficits |
| Urethral Carcinoma | CN 2-12 intact |
| Urethral Carcinoma | intact |
| Urethral Carcinoma | CN 2-12 |
| Urethral Carcinoma | normal creatinine at 0.80 mg/dL |
| Urethral Carcinoma | normal |
| Urethral Carcinoma | creatinine |
| Urethral Carcinoma | distal |
| Urethral Carcinoma | urethral |
| Urethral Carcinoma | lesion |
| Urethral Carcinoma | urethral lesion |
| Urethral Carcinoma | periurethral mass |
| Urethral Carcinoma | invade |
| Urethral Carcinoma | anterior vagina |
| Urethral Carcinoma | no LE edema |
| Urethral Carcinoma | no rash |
| Urethral Carcinoma | No gross neurological deficits, CN 2-12 intact |
| Urethral Carcinoma | normal creatinine |
| Urethral Carcinoma | 0.8 |
| Urethral Carcinoma | distal urethral lesion |
| Urethral Carcinoma | 1.6 x 1.2 x 2.3 |
| Urethral Carcinoma | no evidence of pelvic lymphadenopathy |
| Urethral Carcinoma | no evidence of |
| Urethral Carcinoma | pelvic lymphadenopathy |
| Urethral Carcinoma | pelvic |
| Urethral Carcinoma | lymphadenopathy |
| Urethral Carcinoma | metastatic disease |
| Urethral Carcinoma | metastatic |
| Urethral Carcinoma | disease |
| Urethral Carcinoma | CT urogram |
| Urethral Carcinoma | Pathology from core biopsy |
| Urethral Carcinoma | invasive |
| Urethral Carcinoma | poorly differentiated |
| Urethral Carcinoma | poorly |
| Urethral Carcinoma | carcinoma |
| Urethral Carcinoma | squamous |

| | |
|---|---|
| Urethral Carcinoma | squamous differentiation |
| Urethral Carcinoma | invasive poorly differentiated carcinoma with squamous differentiation |
| Urethral Carcinoma | positive |
| Urethral Carcinoma | CK7 |
| Urethral Carcinoma | p63 |
| Urethral Carcinoma | p16 |
| Urethral Carcinoma | p40 |
| Urethral Carcinoma | GATA3 |
| Urethral Carcinoma | negative |
| Urethral Carcinoma | CK20 |
| Urethral Carcinoma | negative for CK20 |
| Urethral Carcinoma | positive for CK7, p63, p16, p40 and GATA3 |
| NSCLC-IT | BASELINE (*No removals)* |
| NSCLC-IT | former heavy smoker |
| NSCLC-IT | newly diagnosed metastatic carcinoma |
| NSCLC-IT | metastatic carcinoma |
| NSCLC-IT | squamous cell carcinoma from pulmonary origin |
| NSCLC-IT | squamous cell carcinoma |
| NSCLC-IT | pulmonary origin |
| NSCLC-IT | severe left flank pain |
| NSCLC-IT | spontaneous subcapsular renal hematoma |
| NSCLC-IT | left subcapsular hematoma measuring 2.5 cm |
| NSCLC-IT | left subcapsular hematoma |
| NSCLC-IT | right renal cortical cysts measuring 2.5 cm |
| NSCLC-IT | right renal cortical cysts |
| NSCLC-IT | 1.7 cm probable liver cyst |
| NSCLC-IT | liver cyst |
| NSCLC-IT | right 1.6 cm adrenal nodule |
| NSCLC-IT | adrenal nodule |
| NSCLC-IT | exophytic simple cyst in the right kidney |
| NSCLC-IT | exophytic simple cyst |
| NSCLC-IT | heterogenous enhancing lesion in the left upper pole of left kidney with hematoma |
| NSCLC-IT | heterogenous enhancing lesion |
| NSCLC-IT | left upper pole |
| NSCLC-IT | left kidney with hematoma |
| NSCLC-IT | enhancing 2.3 cm lesion in the liver |
| NSCLC-IT | liver |
| NSCLC-IT | right upper lobe mass measuring 4.7 cm |

| | |
|---|---|
| NSCLC-IT | right upper lobe |
| NSCLC-IT | left kidney |
| NSCLC-IT | right renal |
| NSCLC-IT | left subcapsular |
| NSCLC-IT | mediastinal and hilar lymphadenopathy with scattered pulmonary nodules |
| NSCLC-IT | mediastinal |
| NSCLC-IT | hilar |
| NSCLC-IT | lymphadenopathy |
| NSCLC-IT | scattered pulmonary nodules |
| NSCLC-IT | pulmonary nodules |
| NSCLC-IT | liver biopsy |
| NSCLC-IT | poorly differentiated carcinoma |
| NSCLC-IT | 99% |
| NSCLC-IT | 60% |
| NSCLC-IT | CPS 60 |
| NSCLC-IT | TP53, NOTCH2, and NF2 |
| NSCLC-IT | TP53 |
| NSCLC-IT | NOTCH2 |
| NSCLC-IT | NF2 |
| NSCLC-IT | PD-L1 |
| NSCLC-IT | severe right flank pain |
| NSCLC-IT | bilateral subcapsular hematoma in both kidney |
| NSCLC-IT | bilateral |
| NSCLC-IT | both kidney |
| NSCLC-IT | AKI |
| NSCLC-IT | dialysis |
| NSCLC-IT | carcinoma of unknown primary |
| NSCLC-IT | significantly increased uptake in the left tonsils, left lung mass and thoracic lymph nodes, liver |
| NSCLC-IT | significantly increased uptake |
| NSCLC-IT | left tonsils |
| NSCLC-IT | left lung mass |
| NSCLC-IT | thoracic lymph nodes |
| NSCLC-IT | liver |
| NSCLC-IT | left tonsils, left lung mass and thoracic lymph nodes, liver |
| NSCLC-IT | few polyps (tubular adenomas and hyperplastic polyps) |
| NSCLC-IT | few polyps |
| NSCLC-IT | (tubular adenomas and hyperplastic polyps) |
| NSCLC-IT | negative for malignancy |

| | |
|---|---|
| NSCLC-IT | malignancy |
| NSCLC-IT | negative |
| NSCLC-IT | right upper lobe lung mass |
| NSCLC-IT | Positive for non-small cell carcinoma, favoring squamous cell carcinoma (TPS 60%, tumor morphology is similar to that liver biopsy) |
| NSCLC-IT | Positive for non-small cell carcinoma |
| NSCLC-IT | favoring squamous cell carcinoma |
| NSCLC-IT | (TPS 60%, tumor morphology is similar to that liver biopsy) |
| NSCLC-IT | Positive |
| NSCLC-IT | non-small cell carcinoma |
| NSCLC-IT | that liver biopsy |
| NSCLC-IT | 46% |
| NSCLC-IT | pancreatic cancer |
| NSCLC-IT | 46% probability being pancreatic cancer |
| NSCLC-IT | 46% probability being pancreatic cancer and 11% pulmonary squamous cell carcinoma, and 11% probability from head and neck |
| NSCLC-IT | 11% pulmonary squamous cell carcinoma |
| NSCLC-IT | 11% |
| NSCLC-IT | pulmonary squamous cell carcinoma |
| NSCLC-IT | 11% probability |
| NSCLC-IT | 11% probability from head and neck |
| NSCLC-IT | head and neck |
| NSCLC-IT | ambulate without difficulty |
| NSCLC-IT | ambulate |
| NSCLC-IT | difficulty |
| NSCLC-IT | Pulmonary symptoms is stable, without significant worsening of chest pain, dyspnea, hemoptysis |
| NSCLC-IT | Pulmonary symptoms is stable |
| NSCLC-IT | significant worsening of chest pain, dyspnea, hemoptysis |
| NSCLC-IT | chest pain |
| NSCLC-IT | dyspnea |
| NSCLC-IT | hemoptysis |
| NSCLC-IT | urinary and still has indwelling foley catheter |
| NSCLC-IT | urinary |
| NSCLC-IT | indwelling foley catheter |
| NSCLC-IT | any headache, vision changes, hearing changes, dysphagia, nausea, vomiting, abd pain, constipation, diarrhea, dysuria, focal weakness, leg swelling, peripheral neuropathy, fever, chill |
| NSCLC-IT | denies |
| NSCLC-IT | headache |

| | |
|---|---|
| NSCLC-IT | vision changes |
| NSCLC-IT | hearing changes |
| NSCLC-IT | dysphagia |
| NSCLC-IT | nausea |
| NSCLC-IT | vomiting |
| NSCLC-IT | abd pain |
| NSCLC-IT | constipation |
| NSCLC-IT | diarrhea |
| NSCLC-IT | dysuria |
| NSCLC-IT | focal weakness |
| NSCLC-IT | leg swelling |
| NSCLC-IT | peripheral neuropathy |
| NSCLC-IT | fever |
| NSCLC-IT | chill |
| NSCLC-IT | 130/78 |
| NSCLC-IT | (BP Location: Left arm, BP Position: Sitting) |
| NSCLC-IT | 70 |
| NSCLC-IT | 98.2 °F (36.8 °C) (Oral) |
| NSCLC-IT | 18 |
| NSCLC-IT | 1.803 m (5' 11") Comment: reported by pt |
| NSCLC-IT | 100.2 kg (221 lb) Comment: with shoes on |
| NSCLC-IT | 98% |
| NSCLC-IT | 30.82 kg/m¬≤ |
| NSCLC-IT | Heavy Smoker |
| NSCLC-IT | status: 1 |
| NSCLC-IT | male alert and oriented in no acute distress |
| NSCLC-IT | No scleral icterus. EOMI |
| NSCLC-IT | No oropharyngeal lesions or thrush |
| NSCLC-IT | no masses, symmetrical |
| NSCLC-IT | Normal effort. Clear to auscultation: A&P bilaterally, no crackles/rhonchi/wheezes |
| NSCLC-IT | Normal S1/S2, regular rate and rhythm, no murmurs, gallops, rubs. No peripheral edema bilaterally |
| NSCLC-IT | soft, non-tender. Bowel sounds normal |
| NSCLC-IT | no digital cyanosis or clubbing |
| NSCLC-IT | No rashes or lesions |
| NSCLC-IT | Cervical and supraclavicular nodes normal |
| NSCLC-IT | Cranial nerves grossly intact. No focal deficits |
| NSCLC-IT | Affect appropriate for situation. Alert and oriented to person, place and time |

| | |
|---|---|
| NSCLC-IT | Non-small cell carcinoma, favor squamous cell carcinoma |
| NSCLC-IT | Poorly differentiated carcinoma |
| NSCLC-IT | 4.3 CM right upper lobe mass. Scattered pulmonary nodules. Enlarged mediastinal lymph nodes |
| NSCLC-IT | Known progressing pulmonary metastases. Stable subcapsular renal hematoma bilaterally |
| NSCLC-IT | Significantly increased uptake in the left tonsils, left lung mass and thoracic lymph nodes, liver |
| NSCLC-IT | No metastatic disease |
| NSCLC-FU | BASELINE (*No removals)* |
| NSCLC-FU | discoid |
| NSCLC-FU | discoid lupus |
| NSCLC-FU | extensive |
| NSCLC-FU | smoking |
| NSCLC-FU | extensive smoking history |
| NSCLC-FU | history |
| NSCLC-FU | extensive smoking history (40 pack-years, quit 2019) |
| NSCLC-FU | (40 pack-years, quit 2019) |
| NSCLC-FU | squamous cell carcinoma of the lung |
| NSCLC-FU | further management |
| NSCLC-FU | squamous cell carcinoma |
| NSCLC-FU | squamous |
| NSCLC-FU | squamous cell |
| NSCLC-FU | lung |
| NSCLC-FU | hemoptysis |
| NSCLC-FU | LLL completely atelectatic |
| NSCLC-FU | LLL |
| NSCLC-FU | atelectatic |
| NSCLC-FU | completely atelectatic |
| NSCLC-FU | small left pleural effusion |
| NSCLC-FU | small left |
| NSCLC-FU | pleural effusion |
| NSCLC-FU | mildly enlarged AP node |
| NSCLC-FU | small bilateral adrenal lesions (1.5 cm on the right with hounsfield of 7, and 1.3 cm on the left with Hounsfield 20) concern for adenoma |
| NSCLC-FU | small bilateral adrenal lesions |
| NSCLC-FU | (1.5 cm on the right with hounsfield of 7, and 1.3 cm on the left with Hounsfield 20) |
| NSCLC-FU | adenoma |
| NSCLC-FU | fungating bleeding mass |
| NSCLC-FU | left upper lobe |

| | |
|---|---|
| NSCLC-FU | distal left main bronchus |
| NSCLC-FU | Bronch |
| NSCLC-FU | 60-70% obstruction to the airway |
| NSCLC-FU | no abnormal LN station were seen |
| NSCLC-FU | positive |
| NSCLC-FU | Biopsy positive for squamous cell carcinoma |
| NSCLC-FU | arterial embolization |
| NSCLC-FU | initial follow |
| NSCLC-FU | relatively well |
| NSCLC-FU | not had further hemoptysis |
| NSCLC-FU | significant chest pain |
| NSCLC-FU | intermittent mild cough |
| NSCLC-FU | feeling stable |
| NSCLC-FU | Interval History |
| NSCLC-FU | face and chest skin |
| NSCLC-FU | plaquenil |
| NSCLC-FU | denies |
| NSCLC-FU | headache, vision changes, hearing changes, dysphagia, nausea, vomiting, abd pain, constipation, diarrhea, dysuria, focal weakness, leg swelling, peripheral neuropathy, fever, chill |
| NSCLC-FU | headache |
| NSCLC-FU | vision changes |
| NSCLC-FU | hearing changes |
| NSCLC-FU | dysphagia |
| NSCLC-FU | nausea |
| NSCLC-FU | vomiting |
| NSCLC-FU | abd pain |
| NSCLC-FU | constipation |
| NSCLC-FU | diarrhea |
| NSCLC-FU | dysuria |
| NSCLC-FU | focal weakness |
| NSCLC-FU | leg swelling |
| NSCLC-FU | peripheral neuropathy |
| NSCLC-FU | fever |
| NSCLC-FU | chill |
| NSCLC-FU | 160/73 (BP Location: Right arm, BP Position: Sitting) |
| NSCLC-FU | 78 |
| NSCLC-FU | 97.7 °F (36.5 °C) (Oral) |
| NSCLC-FU | 14 |

| | |
|---|---|
| NSCLC-FU | 1.829 m (6') |
| NSCLC-FU | 127 kg (280 lb) Comment: with shoes and jacket on |
| NSCLC-FU | 95% |
| NSCLC-FU | 37.97 kg/m¬≤ |
| NSCLC-FU | Former |
| NSCLC-FU | status: 0 |
| NSCLC-FU | male alert and oriented in no acute distress |
| NSCLC-FU | No scleral icterus. EOMI. |
| NSCLC-FU | No oropharyngeal lesions or thrush |
| NSCLC-FU | no masses, symmetrical |
| NSCLC-FU | Normal effort. Clear to auscultation: A&P bilaterally, no crackles/rhonchi/wheezes |
| NSCLC-FU | Normal S1/S2, regular rate and rhythm, no murmurs, gallops, rubs. No peripheral edema bilaterally |
| NSCLC-FU | soft, non-tender. Bowel sounds normal |
| NSCLC-FU | no digital cyanosis or clubbing |
| NSCLC-FU | No rashes or lesions |
| NSCLC-FU | Cervical and supraclavicular nodes normal |
| NSCLC-FU | Cranial nerves grossly intact. No focal deficits |
| NSCLC-FU | Affect appropriate for situation. Alert and oriented to person, place and time |
| NSCLC-FU | left lower lobe |
| NSCLC-FU | completely |
| NSCLC-FU | abrupt termination of the proximal left lower lobe bronchus |
| NSCLC-FU | obstructive from a mass |
| NSCLC-FU | abrupt termination of the proximal left lower lobe bronchus concern for obstructive from a mass |
| NSCLC-FU | mildly enlarged mediastinal lymph node |
| NSCLC-FU | mediastinal lymph node |
| NSCLC-FU | mildly |
| NSCLC-FU | mildly enlarged |
| NSCLC-FU | enlarged |
| NSCLC-FU | lymph node |
| NSCLC-FU | Small bilateral adrenal lesion |
| NSCLC-FU | adrenal |
| NSCLC-FU | adrenal lesion |
| NSCLC-FU | bilateral |
| NSCLC-FU | Small bilateral |
| NSCLC-FU | None |

Table S2. Sparse autoencoder hyperparameters

| | MedGemma-27B-Text-IT | OpenBioLLM-Llama-70B |
|---|---|---|
| Latents (N) | 65,536 | 98,304 |
| Sparsity (k) | 128 | 128 |
| LLM Layer (L) | 52 | 67 |
| Learning rate | 0.0002 | 0.0002 |
| Auxillary loss sparsity | 512 | 512 |
| Auxillary loss coefficient | 0.03125 | 0.03125 |
| Number steps for reinitializing dead latents | 900 | 900 |
| Optimizer | Adafactor | Adafactor |
| Training steps | 200,000 | 200,000 |
| Batch size (tokens) | 32,768 | 32,768 |
| Validation step size | 2000 | 2000 |
| Clinical Notes | 50,000 | 50,000 |

Table S3. Polysemous medical terms

| **Term** | **Context** |
|---|---|
| Discharge | Hospital departure |
| Discharge | Wound drainage |
| Failure | Cardiac |
| Failure | Renal |
| Failure | Respiratory |
| Block | Cardiac conduction |
| Block | Nerve |
| Block | Anatomical |
| Pressure | Blood pressure |
| Pressure | Intracranial |
| Pressure | Mechanical |
| Lesion | Radiological |
| Lesion | Dermatological |
| Lesion | Pathological |
| Mass | Tumor |
| Mass | Body mass |
| Effusion | Pleural |
| Effusion | Pericardial |
| Effusion | Joint |
| Syndrome | Various clinical syndromes |
| Episode | Cardiac |
| Episode | Neurological |
| Episode | Psychiatric |
| Arrest | Cardiac |
| Arrest | Respiratory |
| Attack | Cardiac |
| Attack | Panic |
| Attack | Transient ischemic |
| Shock | Cardiogenic |

| | |
|---|---|
| Shock | Septic |
| Shock | Hypovolemic |

Table S4. Monosemantic Medical Terms

| **Term** |
|---|
| Hospital |
| Patient |
| Hypertension |
| Treatment |

### SUPPLEMENTARY FIGURES

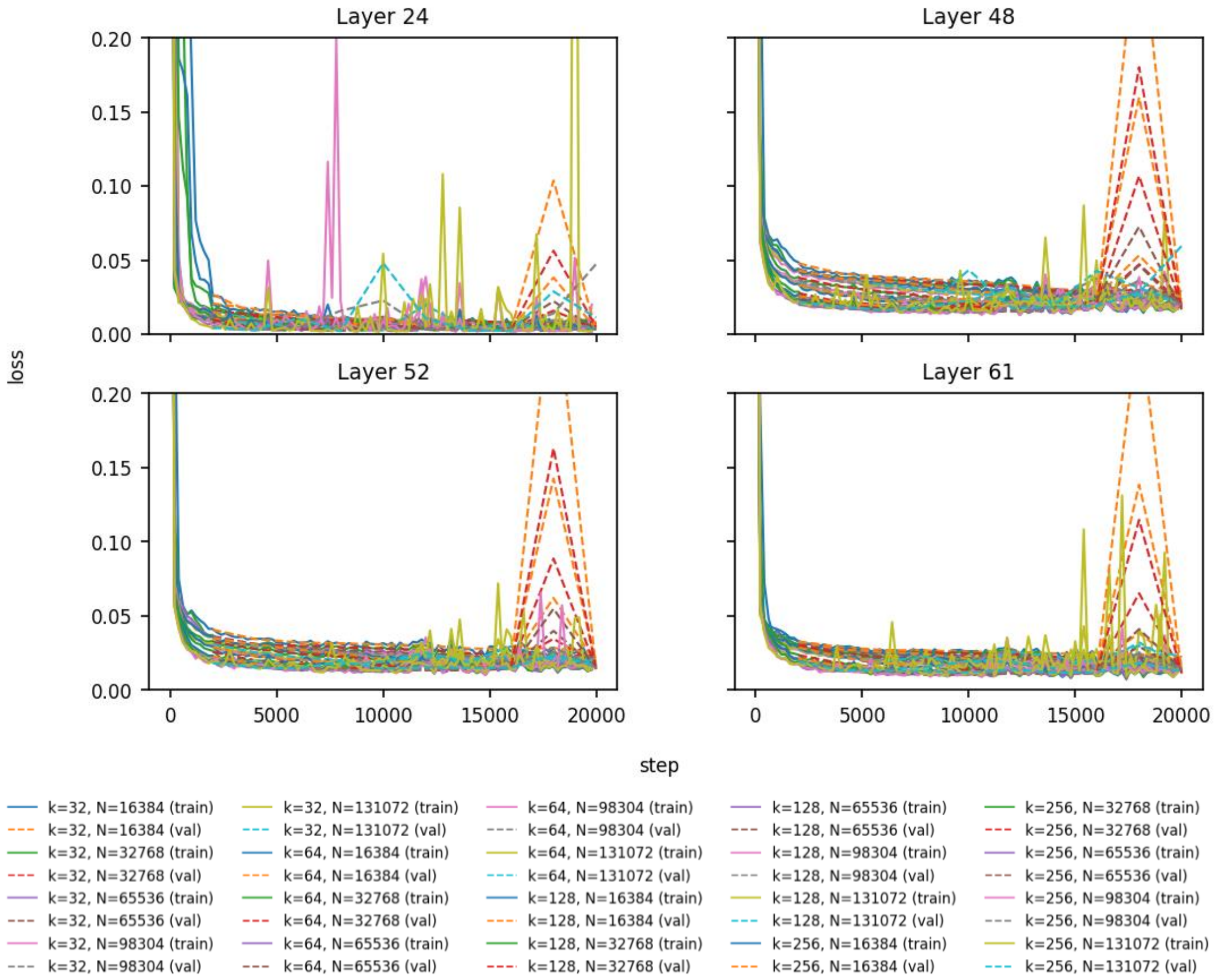

**Fig. S1 | Sparse autoencoder training dynamics across MedGemma layers.** Shown are the normalized mean square error (NMSE) loss curves during SAE training on layers 24 (top left), 48 (top right), 52 (bottom left), and 61 (bottom right) of MedGemma-27B-Text-IT. Each curve represents a different hyperparameter configuration varying sparsity $k \in \{32,64,128,256\}$ and latents $N \in \{16{,}384; 32{,}768; 65{,}536; 98{,}304; 131{,}072\}$. Solid lines indicate training loss while dashed lines represent validation loss. MedGemma SAEs converge to 0.01-0.05 NMSE. Periodic spikes in later training steps indicate learning rate scheduling effects. Loss dynamics are consistent across layers, suggesting similar reconstruction difficulty throughout the network depth.

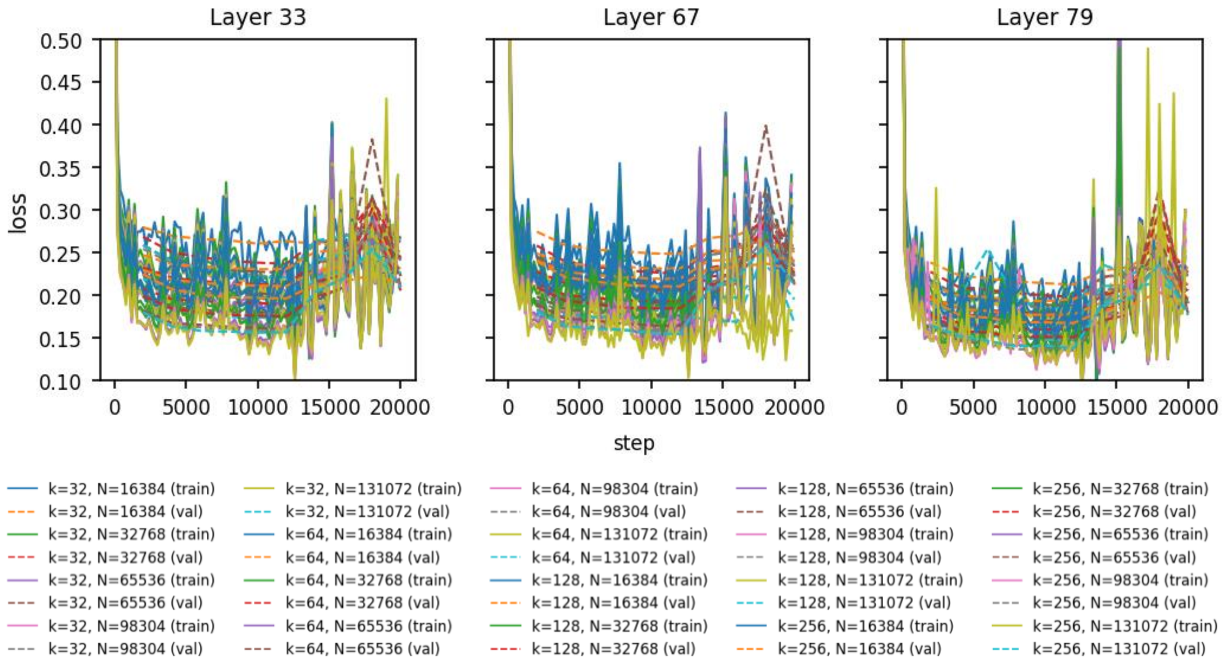

**Fig. S2 | Sparse autoencoder training dynamics across OpenBioLLM layers.** Shown are the normalized mean square error (NMSE) loss curves during SAE training on layers 33 (left), 67 (middle), and 79 (right) of OpenBioLLM-Llama3-70B. Each curve represents a different hyperparameter configuration varying sparsity $k \in \{32,64,128,256\}$ and latents $N \in \{16{,}384; 32{,}768; 65{,}536; 98{,}304; 131{,}072\}$. Solid lines indicate training loss while dashed lines represent validation loss. All configurations converged to a NMSE of 0.15-0.30, with train-validation gaps remaining small indicating minimal overfitting. Periodic spikes in later training steps indicate learning rate scheduling effects. Loss dynamics are consistent across layers, suggesting similar reconstruction difficulty throughout the network depth.

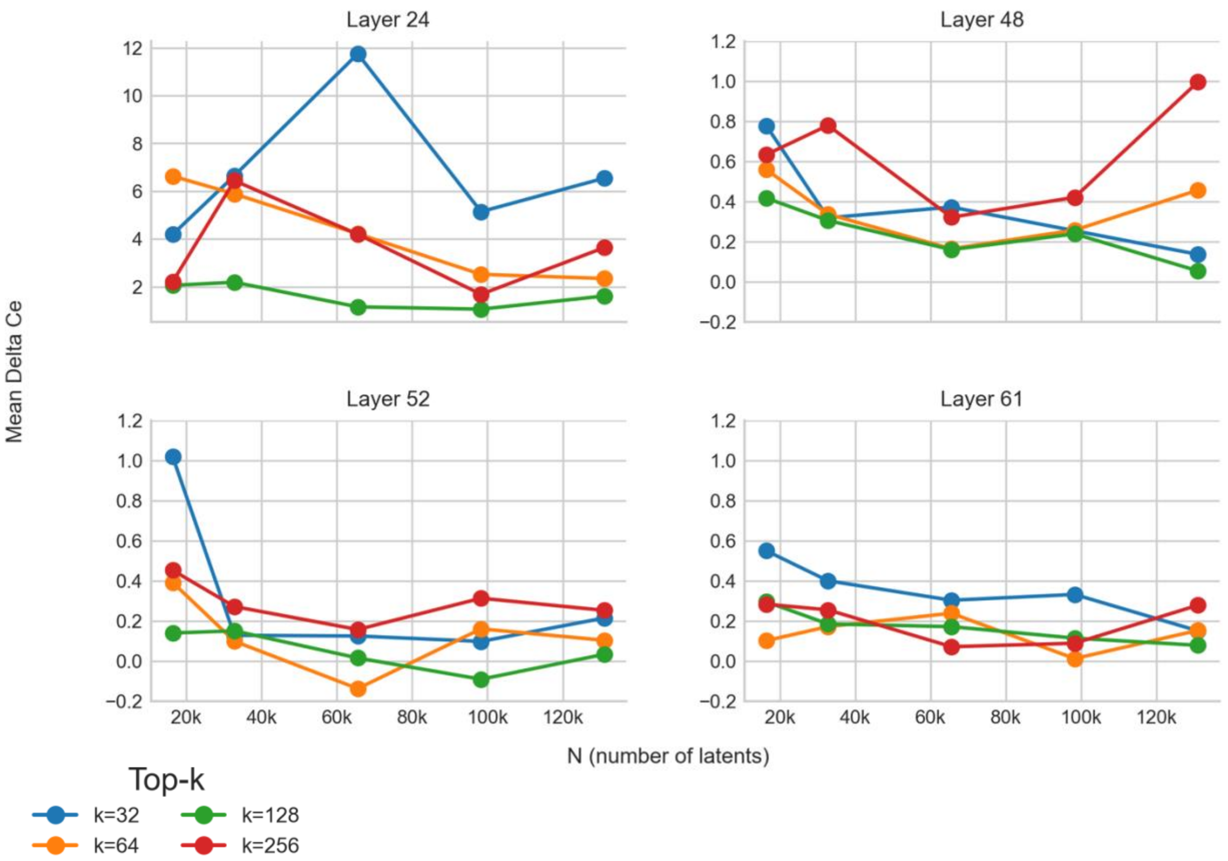

**Fig. S3 | Delta cross-entropy hyperparameter sweep for MedGemma sparse autoencoders.** Shown are the mean delta cross-entropies ($\Delta CE$) which represent how well the sparse autoencoder captures MedGemma-27B-Text-IT's behavior. We sweep across varying sparsity $k \in \{32,64,128,256\}$ and dictionary sizes/latents $N \in \{16,384; 32,768; 65,536; 98,304; 131,072\}$ across layers 24, 48, 52, and 61. Lower $|\Delta CE|$ indicates better preservation of MedGemma's predictive distribution after swapping in the SAE-reconstructed activations. Layer 24 exhibits substantially higher $\Delta CE$ compared to deeper layers (0-12 nats scale versus -0.2-1.2 nats, respectively), reflecting greater reconstruction difficulty in earlier layers. Across layers 48, 52, and 61, k=128 (green) consistently achieves the lowest $|\Delta CE|$, while k=32 shows higher variance and poorer performance, particular at smaller latent sizes. Layer 52 with k=128 and N=65,536 achieves $\Delta CE$ approaching zero, indicating near perfect preservation of model predictions. Thus, we select layer 52, with k=128 and N=65,536, for our downstream interpretability experiments to optimize the tradeoff between sparsity and reconstruction fidelity.

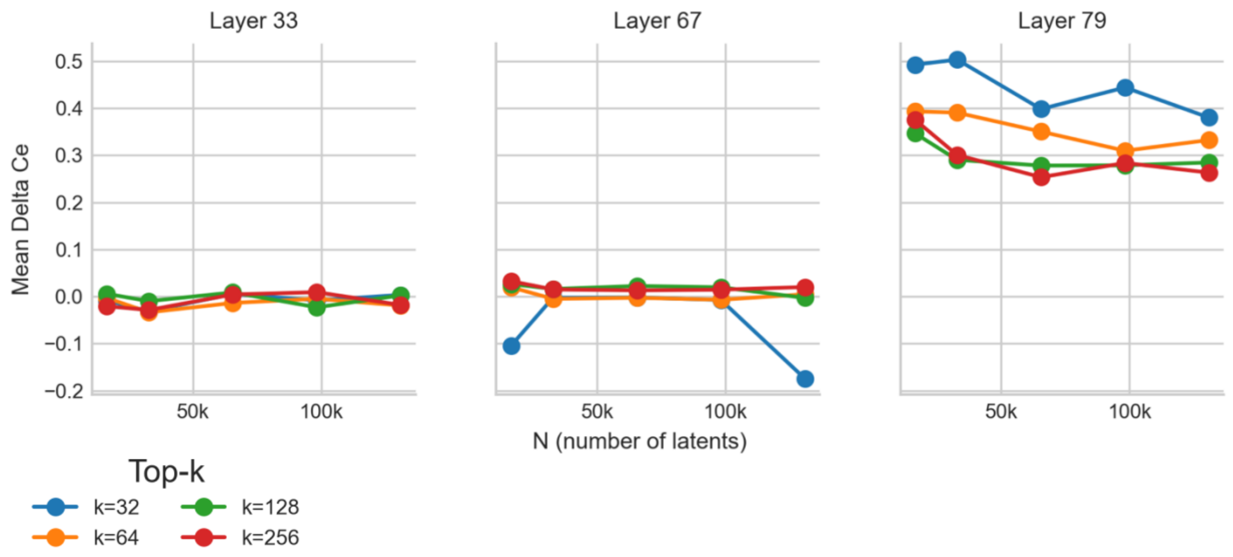

**Fig. S4 | Delta cross-entropy hyperparameter sweep for OpenBioLLM's sparse autoencoders.** Shown are the mean delta cross-entropies ($\Delta CE$) which represent how well the sparse autoencoder captures OpenBioLLM-Llama3-70B's behavior. We sweep across varying sparsity $k \in \{32,64,128,256\}$ and dictionary sizes/latents $N \in \{16{,}384; 32{,}768; 65{,}536; 98{,}304; 131{,}072\}$ across layers 24, 48, 52, and 61. Lower $|\Delta CE|$ indicates better preservation of OpenBioLLM's predictive distribution after swapping in the SAE-reconstructed activations. Layers 33 and 67 achieve near-zero $\Delta CE$ across most configurations, while layer 79 exhibits higher reconstruction fidelity with $\Delta CE$ of 0.25-0.50 nats. Notably, k=32 (blue) produces increasingly negative $\Delta CE$ at layer 67 as dictionary size increases, suggesting unstable reconstruction behavior. In contrast, k=128 (green) maintains $|\Delta CE|$ closest to zero across all layers and dictionary sizes, demonstrating robust preservation of model predictions. We selected k=128 and N=98,304 at layer 67 for downstream analyses since this provides configuration achieves near zero $|\Delta CE|$ while providing a sufficient dictionary capacity (~12x expansion factor) to capture diverse medical concepts without the instability seen for larger dictionary sizes with smaller sparsity.

### SUPPLEMENTARY METHODS (Extended Version)

#### LLM Parameters For Response Generation in Synthetic Oncology Cases

We evaluated three large language models for medical text generation: (i) MedGemma-27B-Text-IT[2], accessed from HuggingFace repository google/medgemma-27b-text-it at commit 6b08c481126ff65a9b8fa5ab4d691b152b8edb5d; (ii) OpenBioLLM-Llama3-70B[3], accessed from HuggingFace repository aaditya/OpenBioLLM-Llama3-70B at commit 7ad17ef0d2185811f731f89d20885b2f99b1e994; and (iii) GPT-5, accessed via the Responses API (model identifier: gpt-5-2025-08-07). All HuggingFace models were loaded in bfloat16 precision using the transformers library. Generation parameters for MedGemma and OpenBioLLM were set to: max_new_tokens=2048, temperature=0.6, top_p=0.9, with self-consistency sampling (n=5). GPT-5 was configured with max_output_tokens=16384, reasoning_effort="medium", and text_verbosity="high".

#### Sparse autoencoder configuration selection

We performed several small-scale experiments to identify the ideal configuration for N (latents), k (sparsity), and L (layer). We train on 5,000 randomly selected clinical notes, with batch size of

4,096 tokens. The training loss curves are shown in **Figs. S1-2** for the sparse autoencoders (SAEs) trained on MedGemma and OpenBioLLM, respectively. **In Fig. S3**, we find that MedGemma's behavior is best captured by the top-k SAE of k=128; N=65,536; and L=52. From **Fig. S4**, for OpenBioLLM, we find the best configuration for behavior is best captured by the top-k SAE of k=128; N=98,304; and L=67. We use these configurations for all further downstream interpretability analyses.
